# Associations of *APOE* e2/e4 dosage and lifestyle on neuroimaging markers in the UK Biobank

**DOI:** 10.1101/2025.05.12.25327420

**Authors:** Yidan Zhang, Dennis van der Meer, Jurjen J. Luykx, David E. J. Linden, Bart P. F. Rutten, Sinan Guloksuz, Gabriella A. M. Blokland

## Abstract

**Background:** Variation in the Apolipoprotein E (APOE) gene is a key genetic determinant of Alzheimer’s disease (AD) risk and is associated with AD-related neuroimaging changes. Lifestyle factors are also associated with AD risk and may interact with APOE to influence neuroimaging biomarkers. However, evidence on these interactions remains limited and inconsistent, particularly regarding composite healthy lifestyle scores in large-scale cohorts.

**Methods:** We analyzed neuroimaging data from 21,014 UK Biobank participants (mean age: 55) to examine neuroimaging markers relevant to AD and assess the independent and interactive effects of *APOE* ε2/ ε4 dosage and healthy lifestyle risk factors. These biomarkers included diffusion tensor imaging (DTI) metrics-fractional anisotropy (FA) and mean diffusivity (MD), white matter hyperintensity (WMH) burden, and volumetric measures of medial temporal lobe regions such as the entorhinal cortex, parahippocampal region, amygdala, and hippocampus.

**Results:** In the *APOE* dosage model, ε4 dosage was associated with lower bilateral hippocampal volume, left parahippocampal volume and lower WM integrity, particularly in the posterior thalamic radiation and cingulum-hippocampus and sagittal stratum. A higher healthy lifestyle score was broadly associated with lower WMH burden and higher WM integrity. No statistically significant evidence was found for effect modification by lifestyle on the association between *APOE* and MRI markers.

**Conclusions:** Our results do not provide support for the idea that healthy lifestyle factors buffer or mitigate the detrimental effects of the APOE dosage on MRI markers, but rather support a primarily outright association between *APOE* ε4 dosage and worse MRI markers.

## Introduction

Alzheimer disease (AD) is a progressive neurodegenerative disease associated with cognitive decline and is the most common form of dementia in the elderly [1]. *APOE* has been known as the single greatest genetic modulator of sporadic late-onset AD risk, with the *ε*4 allele being a well-established genetic risk factor [2]. In Caucasian populations under the age of 65, the most common *APOE* allele is *ε*3 (75%), followed by *ε*4 (15%) and *ε*2 (8%) [3]. The *ε*4 allele is widely considered the ‘risk’ allele due to its association with increased AD risk, while the *ε*3 allele is viewed as neutral, and the *ε*2 allele may have a protective effect. The primary function of the *APOE* gene locus is associated with lipid/cholesterol metabolism, with pleiotropic effects on various biological processes, including neuronal migration, axon guidance, and the clearance of amyloid-*β* plaques in the brain—a hallmark of AD pathology [4].

AD typically develops over decades, with brain changes often manifesting long before dementia symptoms appear [5]. This gradual onset underscores the potential for identifying early biomarkers for preclinical detection and intervention. As a non-invasive imaging tool, magnetic resonance imaging (MRI) enables the identification of these early structural brain changes, potentially signaling the onset of AD pathology. For example, robust volumetric decline is observed in the entorhinal cortex and hippocampus in cases of mild cognitive impairment (MCI) and AD, with changes in the entorhinal cortex often occurring earlier [6,7]. Additionally, diffusion tensor imaging (DTI) metrics, specifically fractional anisotropy (FA) and mean diffusivity (MD), which are commonly used to assess microstructural white matter (WM) integrity, deteriorate as neurodegeneration progresses in AD. Numerous studies indicate that AD risk is associated with WM integrity declines in tracts that connect key gray matter structures involved in memory function, including the parahippocampal WM, the cingulum, the inferior fronto-occipital fasciculus, and the splenium of the corpus callosum [8, 9, 10].

In particular, MRI-detected structural changes in healthy adults carrying different *APOE* genotypes, especially those with higher *ε*4 allele doses, may serve as valuable biomarkers for identifying at-risk individuals before symptoms emerge. Research has shown that in healthy aging, *APOE* ε4 status confers selective vulnerability to volume loss in specific hippocampal subfields [11]. Additionally, an extensive MRI-based study in the UK Biobank (UKB) reported that ε4 carriers exhibit increased white matter hyperintensity (WMH) volume, reflecting greater WM damage, as well as lower WM integrity [12]. Furthermore, in later life, *APOE* ε4 shows a stronger and more consistent association with medial MTL atrophy [2]. However, findings regarding *APOE*’s effects on brain structure have been inconsistent across studies. Some studies have found no significant associations between the ε4 allele and hippocampal volume, overall gray matter, WM volumes, or the integrity of WM tracts [13,14]. One possible explanation for the variability across studies may be age-related differences [15]. *APOE* ε4-related structural changes in MTL also exhibit variability across the lifespan. These inconsistencies may be attributed to methodological heterogeneity, such as variations in DTI metrics and imaging protocols across studies, as well as the potential moderating effects of environmental factors, including healthy lifestyle, which are often not adequately accounted for.

Healthy lifestyle factors—such as a balanced diet, regular physical activity, avoiding smoking, and limiting alcohol consumption—may be linked to brain structure phenotypes relevant to dementia, including gray matter, hippocampal, and WM volumes, as well as WMHs and WM microstructural integrity [16]. While many studies focus on single lifestyle variables, some evidence suggests that combinations of risky lifestyle factors may have cumulative adverse effects on brain structure and function [17]. Essentially, individuals carrying the ε4 allele may be more susceptible to the adverse effects of lifestyle-related risk factors on brain-relevant outcomes. A potential biological explanation is that the APOE locus regulates lipid metabolism, which in turn influences critical brain processes such as WM myelination and neuronal repair. This may render ε4 carriers biologically more vulnerable to the negative impact of suboptimal lifestyle behaviors [18].

Current evidence on the interplay between *APOE* and lifestyle factors remains inconsistent. Most studies have focused on cognitive performance or AD as outcomes. A recent large-scale study using UKB data examined the interaction between APOE ε4 and lifestyle on cognitive function but found no significant interaction effects [19]. Neuroimaging markers may serve as more sensitive indicators of brain changes, yet few studies have examined individual lifestyle factors in this context. One study found that physical activity was associated with better WM integrity in ε4 non-carriers but showed adverse effects in ε4 carriers [20]. Another reported that physical activity (PA) protected hippocampal volume only in those at elevated genetic risk for AD [21]. However, these studies relied on small samples (N < 500), resulting in limited statistical power to detect interaction effects, which may have contributed to inconsistent and potentially spurious findings. Thus far, no large-scale investigation has systematically examined APOE and healthy lifestyle interaction effects on potential neuroimaging biomarkers of AD.

Given the complex roles of APOE and lifestyle behaviors in neurodegenerative processes, this study utilizes the extensive data from the UKB to investigate how APOE and healthy lifestyle independently and interactively influence neuroimaging biomarkers associated with AD risk. Based on prior research, we selected imaging metrics that have been linked to APOE, including DTI measures, total and regional WMH, and volumetric measures within the MTL, including the entorhinal cortex, parahippocampal region, amygdala, and hippocampus. By examining associations between APOE allele dosage/genotype, an integrated healthy lifestyle score, and neuroimaging biomarkers, we aim to provide critical insights into the independent and interactive effects of genetic and healthy lifestyle factors on AD-related neuroimaging alterations. Understanding these interactions is critical for elucidating the pathways through which genetic and lifestyle factors jointly influence AD-related brain changes, and for informing the development of personalized interventions based on modifiable behaviors.

## Methods and Materials

### Study Population

The UKB is a large prospective population-based cohort that includes 502,356 participants with comprehensive phenotypic information. Between 2006 and 2010, all participants attended one of 22 assessment centers, where they underwent a series of physical, sociodemographic, and medical assessments [22]. In 2014, an imaging sub-study was initiated, resulting in neuroimaging data for over 45,000 participants by early 2020 [23]. This study focused on individuals with complete genetic data, lifestyle data, and key MRI outcomes, as well as socioeconomic information. Ethical approval for the UKB was granted by the National Information Governance Board for Health and Social Care and the North West Multicentre Research Ethics Committee (11/NW/0382). All participants provided electronic consent for the use of their anonymized data. This study was conducted under UKB application 55392 using data from the July 2023 release.

### Genetic data

A total of 488,377 UKB participants were genotyped for 805,426 markers using the UK BiLEVE Axiom Array (49,950 participants) and the UKB Axiom Array (438,427 participants). Further information on the genotyping process is available at [http://www.ukbiobank.ac.uk/scientists-3/genetic-data], including detailed technical documentation [https://biobank.ctsu.ox.ac.uk/crystal/ukb/docs/genotyping_sample_workflow.pdf]. We utilized the UKB v3 imputed data, based on the Haplotype Reference Consortium (HRC) reference panel. The BGEN file obtained from UKB was converted to PLINK binary format, and SNPs with more than 10% missingness, as well as those failing the Hardy-Weinberg equilibrium test at *p* < 10^−9^, were filtered out. To remove related individuals up to the third-degree (kinship coefficient > 0.044), kinship coefficients were calculated using KING software based on the provided genotype data. Detailed information on the genotyping process of UKB has been reported elsewhere [24]. The *APOE* genotype was directly determined from two SNPs: rs7412 and rs429358. *APOE* genotype data were analyzed using two approaches: (1) The *APOE* dosage models to assess the quantitative effect of the number of ε2, ε3 and ε4 alleles and (2) a complementary genotype-based analysis using categorical APOE genotypes with ε3/ε3 as the reference.

### Brain MRI acquisition and processing

Brain MRI data were obtained from a subsample of UKB participants who were invited to a follow-up visit that included brain MRI scanning [25,26], the dataset used in the current study consisted of 43,422 participants, all of whom had complete brain MRI scans relevant to our research. Imaging was conducted on 3T Siemens Skyra scanners across three MRI centers, following a publicly available protocol [https://www.fmrib.ox.ac.uk/ukbiobank/protocol/V4_23092014.pdf] and processing pipeline [https://biobank.ctsu.ox.ac.uk/crystal/crystal/docs/brain_mri.pdf]. Details regarding imaging-derived phenotypes and processing are described elsewhere [27]. Structural MRI outcomes derived from T1-weighted images included subcortical volumes and specific regions within the MTL: entorhinal cortex, parahippocampus, amygdala, and hippocampus. Total WMH volumes were calculated based on T1 and T2 FLAIR, derived by UKB using the Brain Intensity Abnormality Classification Algorithm (BIANCA) [28] with the procedure detailed by Miller et al [29]. Periventricular WMH (PWMH) and deep WMH (DWMH) volumes, defined as being less or more than 10 mm distant from the lateral ventricles, respectively [30], were extracted as subphenotypes of WMHs. For diffusion MRI scans were processed using the Tract-Based Spatial Statistics (TBSS) pipeline in the FMRIB Software Library (FSL) [https://biobank.ctsu.ox.ac.uk/crystal/crystal/docs/brain_mri.pdf]. WM tracts were defined using 48 standard-space tract masks developed by Mori et al. at Johns Hopkins University [31]. FA quantifies the directional coherence of water diffusion, while MD reflects the rotationally invariant average diffusivity within brain tissue, both serving as indicators of WM integrity. A full list of selected sMRI and DTI phenotypes is provided in Supplementary Table 1.

### Healthy Lifestyle Factors

Drawing on the healthy lifestyle score used in a previous study [32], based on American Heart Association guidelines [33], we constructed a similar score incorporating four factors: non-smoking status, regular physical activity, healthy diet, and non-daily alcohol use. Each item was scored 0 or 1, yielding a total lifestyle score between 0 (least healthy) to 4 (most healthy). Lifestyle factors were derived from self-reported data collected via the touchscreen questionnaire at baseline recruitment. Non-smoking status was defined as not currently smoking. Non-daily alcohol use was defined as consuming alcohol less frequently than ‘daily or almost daily’. Regular physical activity was defined as ≥150 minutes of moderate or ≥75 minutes of vigorous activity per week. The healthy diet component was defined based on prior dementia-related studies [34,35] and cardiometabolic dietary recommendations [36], which are associated with better late-life cognition and reduced dementia risk [37]. It was assessed using a seven-item score, where meeting each of the following criteria was awarded 1 point: 1) Fruits (≥3 servings/day), 2) Vegetables (≥3 servings/day), 3) Fish (≥2 servings/week), 4) Processed meats (≤2 serving/week), 5) Unprocessed red meats (≤1.5 servings/week), 6) Whole grains (≥3 servings/day), and 7) Refined grains (≤1.5 servings/day). A total score greater than 4 was classified as a healthy diet and coded as 1, while scores of 4 or below were coded as 0. Details on the UKB database fields used for lifestyle indicators and diet are provided in **Supplementary Table 2 and Table 3**.

### Covariates

Baseline assessments gathered information on educational attainment using a touch-screen questionnaire, which was regrouped according to previous studies [38], educational qualifications were categorized into four groups reflecting similar years of education: (1) College/University Degree; (2) Education to age 18 or above, but not at degree level (e.g., General Certificate of Education Advanced Level [A levels] / Advanced Subsidiary Level [AS levels], National Vocational Qualification [NVQ], Higher National Diploma [HND], Higher National Certificate [HNC], or equivalent professional qualifications); (3) Education to age 16 (e.g., General Certificate of Education Ordinary Level [O levels] / General Certificate of Secondary Education [GCSEs], Certificate of Secondary Education [CSEs] or equivalent); and (4) No qualifications. Household income before tax was categorized into five groups: <£18,000, £18,000–£29,999, £30,000–£51,999, £52,000–£100,000, and >£100,000. Employment status was categorized into three groups: (1) Paid employment or self-employed, (2) Retired, and (3) Other unemployment, which included looking after home and/or family, being unable to work due to sickness or disability, being unemployed, doing unpaid or voluntary work, or being a full- or part-time student. The time elapsed between the baseline assessment and the neuroimaging visit (in years) was included as a covariate to account for the interval between data collection and MRI measurements. Intracranial volume (ICV) was included to adjust for individual differences in brain size in analyses involving structural MRI outcomes.

### Statistical Analysis

Participants who reported a neurological condition at baseline or during the scanning visit (∼ 5%; see **Supplementary Table 4**) were excluded, following the exclusion criteria outlined in a previous study [14]. Individuals with rare *APOE* genotypes (e.g., ε1/ε4) were also excluded. Further exclusions included non-White British participants, third-degree or closer relatives (based on kinship coefficients), and those with MRI data outliers beyond ±4 SD. Covariates and MRI phenotypes were stratified by *APOE* genotype for descriptive analysis.

Spearman’s rank correlation were used to assess the potential gene–environment correlation between *APOE* dosage and the total healthy lifestyle score to rule out gene– environment correlation that may confound interaction analysis.

In primary analyses, linear regression models were used to assess: (i) the dose-dependent associations of *APOE* ε4, ε2, and *APOE* genotype (ε2/ε2, ε2/ε3, ε2/ε4, ε3/ε4, ε4/ε4 with ε3/ε3 as the reference) separately with each neuroimaging outcome. *APOE* ε4 dosage effects were evaluated in a subsample excluding *APOE* ε2 carriers to avoid potential confounding by ε2’s protective influence (the inverse holds for *APOE* ε2 dosage effects); (ii) the associations of lifestyle factors with neuroimaging outcomes; and (iii) the interactions between *APOE* ε4, ε2, and *APOE* genotype and lifestyle factors were assessed using both continuous interaction terms (e.g., *APOE* dosage × total lifestyle score) and categorical interaction terms (e.g., *APOE* carrier status × lifestyle category); (iv) the potential moderation of these associations by sex or age. In the categorical interaction models, *APOE* dosage was classified as carrier (1–2 alleles) vs. non-carrier (0 alleles), and total lifestyle score (TLS) was categorized as Healthy (score = 4), Intermediate (score = 3), and Unhealthy (score = 0–2).

All primary analyses were based on fully adjusted models, including baseline age, age², sex, imaging assessment center, time elapsed from baseline to the neuroimaging visit (in years), ICV for volumetric outcomes, educational attainment, employment status, household income, long-standing illness, and 20 genetic principal components for *APOE-*related analyses to account for ancestry subpopulation structure.

In sensitivity analyses, we conducted the following:(i) repeated the models using a partially adjusted covariate set (excluding socioeconomic and health status variables) to evaluate the robustness of the findings; (ii) tested whether the *APOE* × lifestyle interaction was robust to an alternative grouping of the TLS, by comparing a binary categorization (Healthy: score = 3-4 vs. Non-healthy: score = 0–2) to the original three-level classification; (iii). assessed the interaction between *APOE* and dietary score (0–7), the only available individual lifestyle component with a quantitative value, on neuroimaging outcomes.

All WMH volumes were log-transformed to correct for skewness, and all brain MRI metrics were standardized into Z-scores. Power calculations were conducted using G*Power 3 [39] to confirm that the available sample size was sufficient for the planned analyses. FDR correction was applied within each MRI modality category (DTI, volume, WMH) across all regions [40], which is consistent with prior neuroimaging studies analyzing distinct imaging modalities [41].

All statistical analyses were performed using R version 4.2.2 (https://www.r-project.org/).

## Results

### Descriptive Statistics

After applying the exclusions mentioned above, the analyzed sample comprised 21,014 UKB participants (**Supplementary Figure 1**). It was determined that the study had 99% power to detect a small effect size (Cohen’s *f*^2^ = 0.01) for significant associations and interactions with a sample size of n=4,585, suggesting the current analyses have generally good power. Compared to the excluded participants, the included sample was younger, healthier, and socioeconomically advantaged, as evidenced by higher educational attainment, higher house income, higher employment rates, less long-standing illness, and healthier lifestyles (all p<0.05) (**Supplementary Table 5**). **Table 1** summarizes the baseline characteristics of participants included in the analysis, both overall and stratified by *APOE* genotype. The frequencies of the *ε*2, *ε*3, and *ε*4 alleles were 8.0%, 76.8%, and 15.2%, respectively. Furthermore, 75.5% of the participants had a favorable lifestyle score (≥3). Small significant differences in age were observed across genotypes(p<0.001), while other sociodemographic characteristics showed no significant variation by *APOE* genotype. The FA and MD metrics for specific WM tracts, both overall and categorized by *APOE* genotype are displayed in **Supplementary Table 6**. Significant genotype-based differences were identified in the Cingulum (hippocampus) for both FA and MD, with the most pronounced effects observed at p < 0.001.

**Table 1.** Baseline Characteristics of Participants by *APOE* Genotype.

| Characteristic | Overall<br>N = 21,014<sup>1</sup> | e3e3<br>N = 12,355<sup>1</sup> | e2e2<br>N = 120<sup>1</sup> | e2e3<br>N = 2,638<sup>1</sup> | e2e4<br>N = 499<sup>1</sup> | e3e4<br>N = 4,932<sup>1</sup> | e4e4<br>N = 470<sup>1</sup> | p-value<sup>2</sup> |
| --- | --- | --- | --- | --- | --- | --- | --- | --- |
| Age | 54.596 (7.488) | 54.765 | 53.983 | 54.804 | 54.315 | 54.178 (7.425) | 53.821 | <0.001 |
| Sex |  |  |  |  |  |  |  | 0.267 |
| Female | 10,507 (50%) | 6,111 (49%) | 62 (52%) | 1,324 | 266 (53%) | 2,494 (51%) | 250 (53%) | - |
| Male | 10,507 (50%) | 6,244 (51%) | 58 (48%) | 1,314 | 233 (47%) | 2,438 (49%) | 220 (47%) | - |
| Education |  |  |  |  |  |  |  | 0.279 |
| College/University | 10,214 (49%) | 6,004 (49%) | 57 (48%) | 1,275 | 256 (51%) | 2,381 (48%) | 241 (51%) | - |
| Education to age 16 | 4,833 (23%) | 2,811 (23%) | 30 (25%) | 592 (22%) | 120 (24%) | 1,168 (24%) | 112 (24%) | - |
| Education to age 18 | 4,833 (23%) | 2,855 (23%) | 27 (23%) | 620 (24%) | 95 (19%) | 1,149 (23%) | 87 (19%) | - |
| No qualifications | 1,134 (5.4%) | 685 (5.5%) | 6 (5.0%) | 151 (5.7%) | 28 (5.6%) | 234 (4.7%) | 30 (6.4%) | - |
| Income Status |  |  |  |  |  |  |  | 0.195 |
| < £18,000 | 2,220 (11%) | 1,299 (11%) | 10 (8.3%) | 278 (11%) | 64 (13%) | 523 (11%) | 46 (9.8%) | - |
| £18,000–£30,999 | 4,498 (21%) | 2,716 (22%) | 17 (14%) | 578 (22%) | 102 (20%) | 997 (20%) | 88 (19%) | - |
| £31,000–£51,999 | 6,322 (30%) | 3,676 (30%) | 42 (35%) | 795 (30%) | 149 (30%) | 1,513 (31%) | 147 (31%) | - |
| £52,000–£100,000 | 6,210 (30%) | 3,617 (29%) | 37 (31%) | 799 (30%) | 142 (28%) | 1,466 (30%) | 149 (32%) | - |
| > £100,000 | 1,764 (8.4%) | 1,047 | 14 (12%) | 188 (7.1%) | 42 (8.4%) | 433 (8.8%) | 40 (8.5%) | - |
| Employment Status |  |  |  |  |  |  |  | 0.089 |
| Employed | 14,854 (71%) | 8,669 (70%) | 79 (66%) | 1,854 | 368 (74%) | 3,530 (72%) | 354 (75%) | - |
| Retired | 5,152 (25%) | 3,099 (25%) | 31 (26%) | 659 (25%) | 108 (22%) | 1,161 (24%) | 94 (20%) | - |
| Unemployed | 1,008 (4.8%) | 587 (4.8%) | 10 (8.3%) | 125 (4.7%) | 23 (4.6%) | 241 (4.9%) | 22 (4.7%) | - |
| Longstanding Illness |  |  |  |  |  |  |  | 0.228 |
| No | 16,691 (79%) | 9,792 (79%) | 98 (82%) | 2,077 | 402 (81%) | 3,963 (80%) | 359 (76%) | - |
| Yes | 4,323 (21%) | 2,563 (21%) | 22 (18%) | 561 (21%) | 97 (19%) | 969 (20%) | 111 (24%) | - |
| Alcohol Frequency |  |  |  |  |  |  |  | 0.596 |
| High Frequency | 4,825 (23%) | 2,840 (23%) | 31 (26%) | 587 (22%) | 104 (21%) | 1,146 (23%) | 117 (25%) | - |
| Low Frequency | 16,189 (77%) | 9,515 (77%) | 89 (74%) | 2,051 | 395 (79%) | 3,786 (77%) | 353 (75%) | - |
| Smoking Status |  |  |  |  |  |  |  | 0.654 |
| Current smoker | 1,145 (5.4%) | 667 (5.4%) | 3 (2.5%) | 138 (5.2%) | 29 (5.8%) | 283 (5.7%) | 25 (5.3%) | - |
| Non-smoker | 19,869 (95%) | 11,688 | 117 (98%) | 2,500 | 470 (94%) | 4,649 (94%) | 445 (95%) | - |
| Healthy Diet |  |  |  |  |  |  |  | 0.279 |
| Unhealthy | 9,888 (47%) | 5,786 (47%) | 56 (47%) | 1,290 | 248 (50%) | 2,295 (47%) | 213 (45%) | - |
| Healthy | 11,126 (53%) | 6,569 (53%) | 64 (53%) | 1,348 | 251 (50%) | 2,637 (53%) | 257 (55%) | - |
| Physical Activity |  |  |  |  |  |  |  | 0.286 |
| No | 4,425 (21%) | 2,558 (21%) | 21 (18%) | 565 (21%) | 113 (23%) | 1,053 (21%) | 115 (24%) | - |
| Yes | 16,589 (79%) | 9,797 (79%) | 99 (83%) | 2,073 | 386 (77%) | 3,879 (79%) | 355 (76%) | - |
| Total Healthy Lifestyle |  |  |  |  |  |  |  | 0.984 |
| 0 | 50 (0.2%) | 31 (0.3%) | 0 (0%) | 7 (0.3%) | 1 (0.2%) | 11 (0.2%) | 0 (0%) | - |
| 1 | 807 (3.8%) | 467 (3.8%) | 3 (2.5%) | 105 (4.0%) | 18 (3.6%) | 193 (3.9%) | 21 (4.5%) | - |
| 2 | 4,289 (20%) | 2,487 (20%) | 21 (18%) | 541 (21%) | 108 (22%) | 1,028 (21%) | 104 (22%) | - |
| 3 | 9,084 (43%) | 5,352 (43%) | 60 (50%) | 1,155 | 220 (44%) | 2,098 (43%) | 199 (42%) | - |
| 4 | 6,784 (32%) | 4,018 (33%) | 36 (30%) | 830 (31%) | 152 (30%) | 1,602 (32%) | 146 (31%) | - |
| Baseline to MRI | 9.283 (1.991) | 9.262 | 9.079 | 9.310 | 9.269 | 9.334 (2.010) | 9.194 | 0.105 |
| Entorhinal Volume | 3,517.751 | 3,521.517 | 3,531.192 | 3,519.353 | 3,478.643 | 3,516.470 | 3,461.323 | 0.116 |
| Parahippocampal | 4,197.683 | 4,204.667 | 4,167.425 | 4,183.360 | 4,205.212 | 4,190.344 | 4,171.260 | 0.131 |
| Amygdala Volume | 3,303.740 | 3,305.294 | 3,282.840 | 3,297.584 | 3,293.089 | 3,309.330 | 3,255.432 | 0.184 |
| Hippocampus Volume | 8,095.636 | 8,103.759 | 8,040.104 | 8,083.115 | 8,070.508 | 8,095.927 | 7,990.183 | 0.110 |
| WMH from T1/T2 | 7.942 (0.950) | 7.953 | 7.845 | 7.917 | 7.843 | 7.937 (0.949) | 7.962 | 0.098 |
| Periventricular WMH | 7.748 (0.935) | 7.761 | 7.645 | 7.722 | 7.646 | 7.744 (0.934) | 7.737 | 0.061 |
| Deep WMH (mm<sup>3</sup>) | 5.923 (1.311) | 5.932 | 5.860 | 5.907 | 5.829 | 5.906 (1.313) | 6.066 | 0.054 |
| gFA (AU) | 0.563 (0.018) | 0.564 | 0.564 | 0.564 | 0.563 | 0.563 (0.018) | 0.562 | 0.454 |
| gMD (AU) | 0.001 (0.000) | 0.001 | 0.001 | 0.001 | 0.001 | 0.001 (0.000) | 0.001 | 0.153 |

<sup>1</sup> Mean (SD); n (%)
<sup>2</sup> Kruskal-Wallis rank sum test; Pearson's Chi-squared test with simulated *p*-value (based on 2000 replicates); Fisher's exact test for count data with simulated *p*-value (based on 2000 replicates).
Abbreviations: gFA, global fractional anisotropy; gMD, global mean diffusivity; AU, arbitrary units.

### Association of *APOE* dosage (ε2/ε4) with MRI markers

We examined the associations between *APOE* ε4 dosage and all MRI markers within the subset excluding ε2 allele carriers (i.e., excluding ε2/ε2, ε2/ε3, and ε2/ε4 genotypes; n = 17,757).

For DTI measures (**Figure 1**), higher *APOE* ε4 dosage was significantly associated with reduced WM integrity (lower FA and higher MD). For FA, ε4 dosage showed significant negative associations, with in order of effect size, Posterior Thalamic Radiation (right PTR: β [95% CI] = -0.050 [-0.074, -0.026], *p*_FDR_ = 0.001), Cingulum Hippocampus (Cing_HIP; bilateral), fornix/stria terminalis (right; fornix ST), Middle Cerebellar Peduncle (right; MCP), and sagittal stratum (right, SStratum). For MD, the significant tracts, ordered by effect size, involved bilateral PTR (right PTR: β [95% CI] = 0.056 [0.032, 0.080], *p*_FDR_ < 0.001), Cing_HIP (bilateral), SStratum (bilateral), Posterior Limb of Internal Capsule (PLIC), Genu of Corpus Callosum (GenuCC) and Splenium of Corpus Callosum (bilateral; splCC), Superior Longitudinal Fasciculus (SLF), Anterior Limb of Internal Capsule (ALIC), SCR, and Body of Corpus Callosum (BodyCC). Corresponding forest plots for APOE associations with FA and MD are available in **Supplementary Figures 2 and 3**.

**Figure 1.**
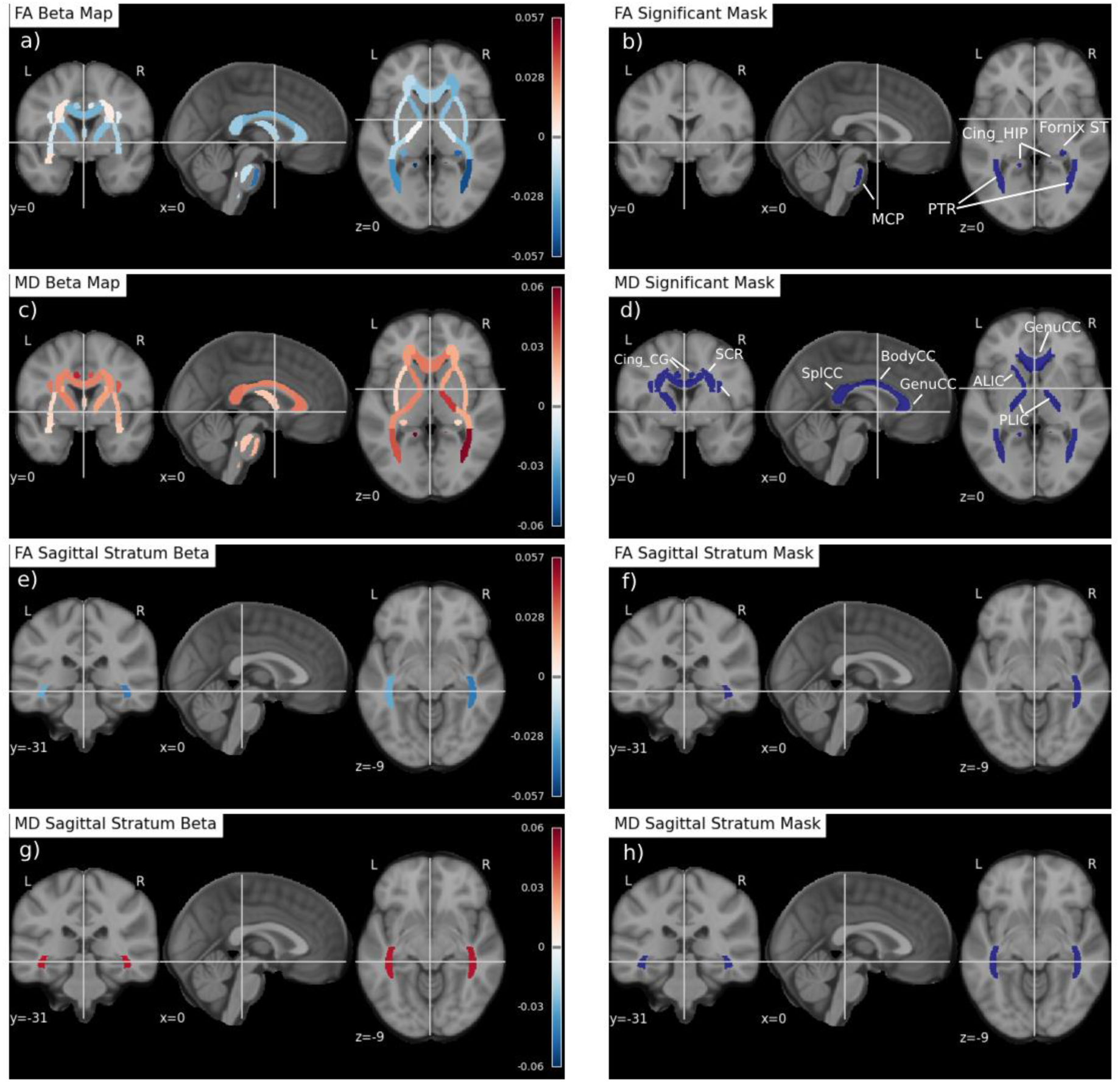
Brain maps showing significant associations between APOE ε4 and DTI measures (FA/MD). **a)** Whole-brain FA results visualized using standardized beta values (color intensity indicates effect size); **b)** Tracts showing statistically significant associations between APOE ε4 dosage and fractional anisotropy (FA); **c)** Whole-brain MD results visualized using standardized beta values; **d)** Tracts showing statistically significant associations between APOE ε4 dosage and mean diffusivity (MD); **e–h)** The sagittal stratum, which showed significant associations with APOE ε4 in both FA and MD analyses, is visualized separately due to its limited visibility in the whole-brain maps. Cut coordinates (x = 0, y = 31, z = 9) were selected to optimally display this tract. ***Abbreviations*** *are listed at the end of the manuscript*.

For volumes of MTL structures and WMHs (**Figure 2**), we found that APOE ε4 dosage was associated with smaller hippocampal volume (total and bilateral) and smaller parahippocampal volume (total and right). No associations were found with WMH measures (all *p*_FDR_ > 0.05).

**Figure 2.**
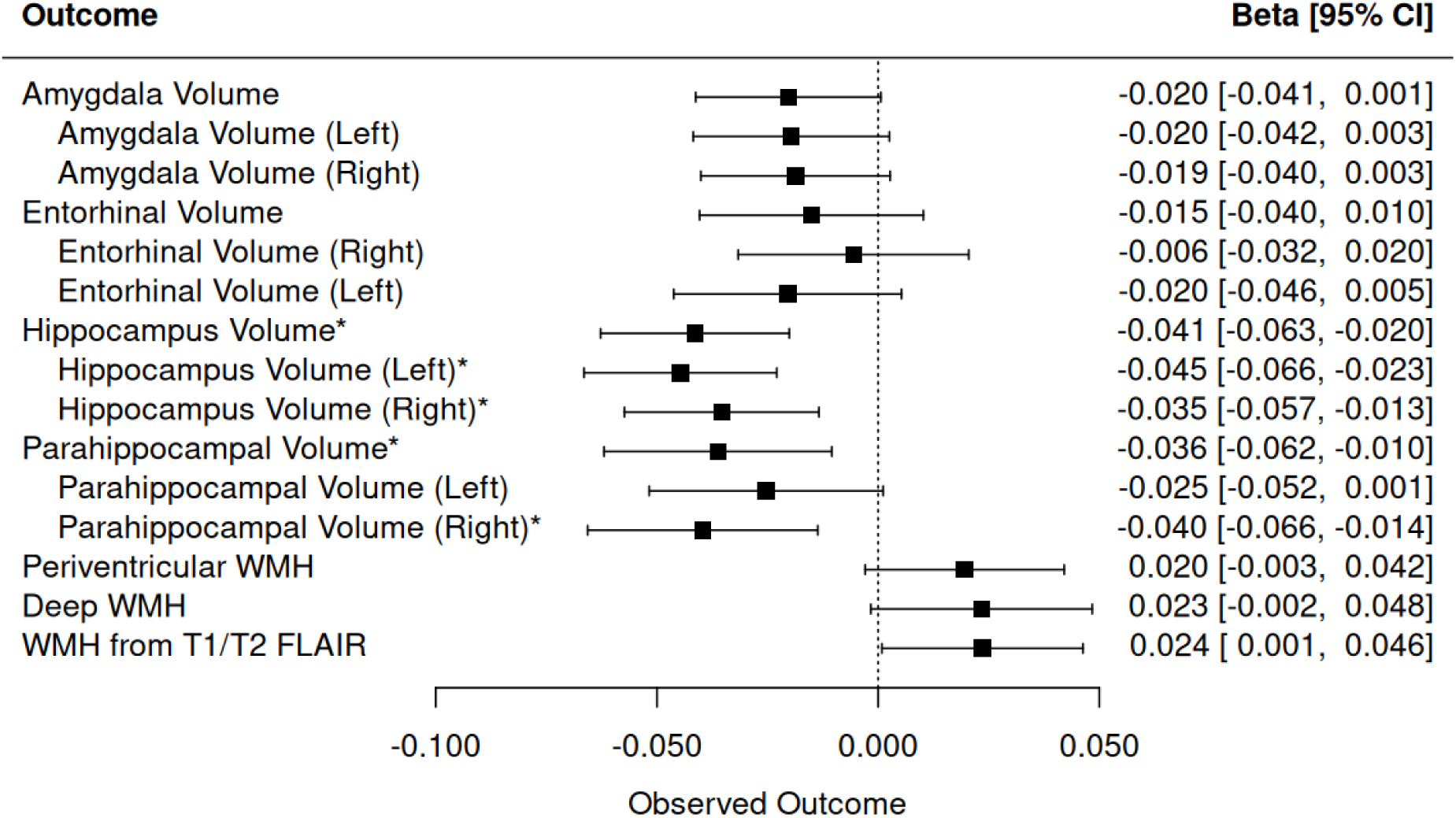
Forest plot of the associations between APOE ε4 dosage and medial temporal lobe subregional volumes and white matter hyperintensities. ***Abbreviations*** *are listed at the end of the manuscript*.

Subsequently, the associations between APOE ε2 dosage and all MRI markers were examined for the subset excluding ε4 allele carriers (i.e., excluding ε4/ε4, ε4/ε3, ε4/ε2 genotypes; n = 15,113). No significant ε2 dosage effects were observed across any MRI measures (all *p*_FDR_ > 0.05).

Full results of *APOE* dosage (ε2 and ε4) associations with MRI markers—including all tracts, standardized β coefficients, standard errors, 95% confidence intervals, R² values, and both uncorrected and FDR-adjusted p-values—are presented in **Supplementary Table 7**.

No significant interactions were found between *APOE* allele dosage and age on MRI outcomes (**Supplementary Table 8**). However, when testing for interactions with sex, one significant result was observed for *APOE* ε2 dosage on FA in the right tapetum (β interaction= 0.128, 95% CI = [0.057, 0.198], *p*_FDR_ = 0.036), suggesting a potentially more protective effect of ε2 in males than in females on this MRI marker, with no similar FDR significant interaction observed for other MRI measures. (see **Supplementary Table 9**).

#### APOE genotypes

Significant associations were identified for ε4/ε4 carriers, who exhibited the greatest differences relative to ε3/ε3 carriers. For the WM Integrity, APOE ε4/ε4 carriers demonstrated significantly lower WM integrity in PTR, SStratum, Cing_HIP, Cingulum Cingulate Gyrus, SLF, and PLIC. For the WMH, deep WMH burden was significantly greater in APOE ε4/ε4 carriers compared to ε3/ε3 carriers, with an increase of 0.14 SD (β [95% CI] = 0.137 [0.056, 0.219], p = 9.9×10⁻⁴, *p*_FDR_ = 0.015).For MTL volume changes, bilateral hippocampal and amygdala volumes were significantly smaller in APOE ε4/ε4 carriers compared to ε3/ε3 carriers, with effect sizes ranging from -0.102 to -0.167 SDs. Compared to the dosage model, the WM tracts identified here were largely contained within those associated with ε4 dosage. Additionally, deep WMH was detected in the genotype analysis but not in the dosage model. Hippocampal involvement was consistently detected across both analyses; however, associations shifted from the parahippocampal region in the dosage model to the amygdala in the genotype analysis (**Figure 3**; full results in **Supplementary Table 10**).

**Figure 3.**
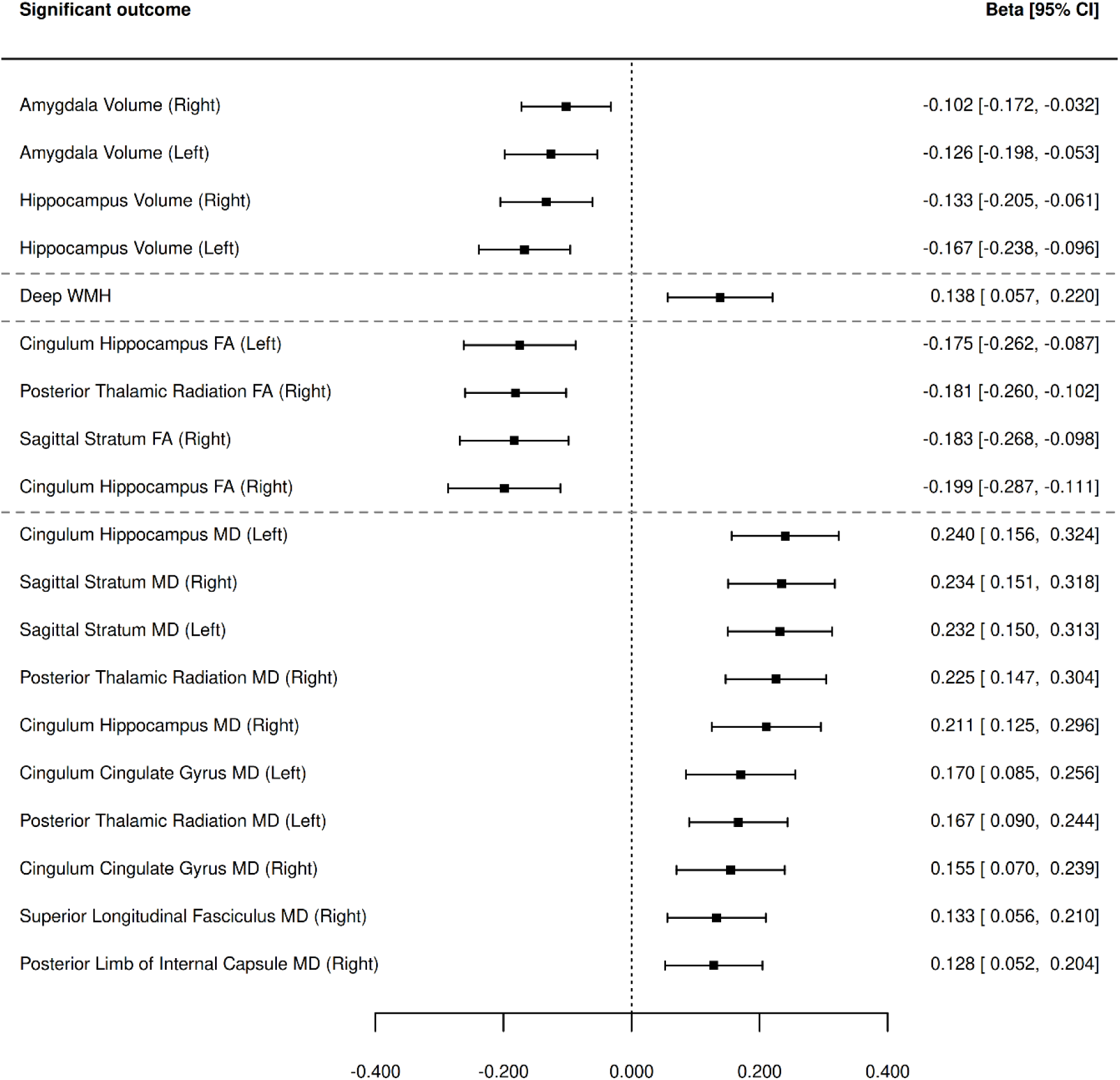
Forest plot of the significant associations between APOE ε4/ε4 (compared to ε3/ε3) and MRI markers. Genotypes e2/e2, e2/e3, e2/e4, e3/e4, and e4/e4 were compared against the neutral-risk reference group (e3/e3). Only the e4/e4 genotype showed a statistically significant association with MRI markers relative to e3/e3. Results shown reflect only those associations that survived multiple comparison correction (FDR-adjusted p < 0.05).

### Association of healthy lifestyle factors with MRI markers

For DTI measures, a healthier lifestyle as indexed by the THLS score was associated with higher FA and lower MD across multiple tracts, particularly in periventricular WM regions (e.g., ALIC, PLIC, SCR (Superior Corona Radiata), Anterior Corona Radiata (ACR), BodyCC, GenuCC and the fornix. A few associations were also found in deeper WM tracts, such as the Superior Fronto-Occipital Fasciculus (SFOF). Higher total healthy lifestyle scores were significantly associated with a lower burden of total WMH (β [95% CI]= -0.020 [-0.034, -0.007], *p*_FDR_ = 7.93 × 10⁻³), and a similar association was observed for periventricular WMH volume. However, the total healthy lifestyle score was not significantly associated with any MTL subcortical volumes. Overall, the pattern suggests widespread benefits of a healthier lifestyle on WM microstructure (**Figure4a**).

Among the four individual lifestyle factors, smoking status and frequency of alcohol intake showed the strongest associations across MRI measures. For illustration, non-smoking was most strongly associated with lower WMH burden (β [95% CI] = -0.172 [-0.220, -0.124], *p*_FDR_ < 0.001) and higher white matter integrity across several major tracts (|β| range: 0.065– 0.173, all *p*_FDR_ < 0.05). Similarly, lower frequency of alcohol intake was associated with higher white matter integrity, most notably in the genu of the corpus callosum (FA: β [95% CI] = 0.100 [0.071, 0.127], *p*_FDR_ < 0.001), and with larger volumes of the left amygdala and left hippocampus (β = 0.047, p = 0.006 for both). The other lifestyle factors, including physical activity and healthy diet, also showed favorable associations with WM integrity (**Figure 4b–e**). Comprehensive results are presented in **Supplementary Table 11**.

**Figure 4.**
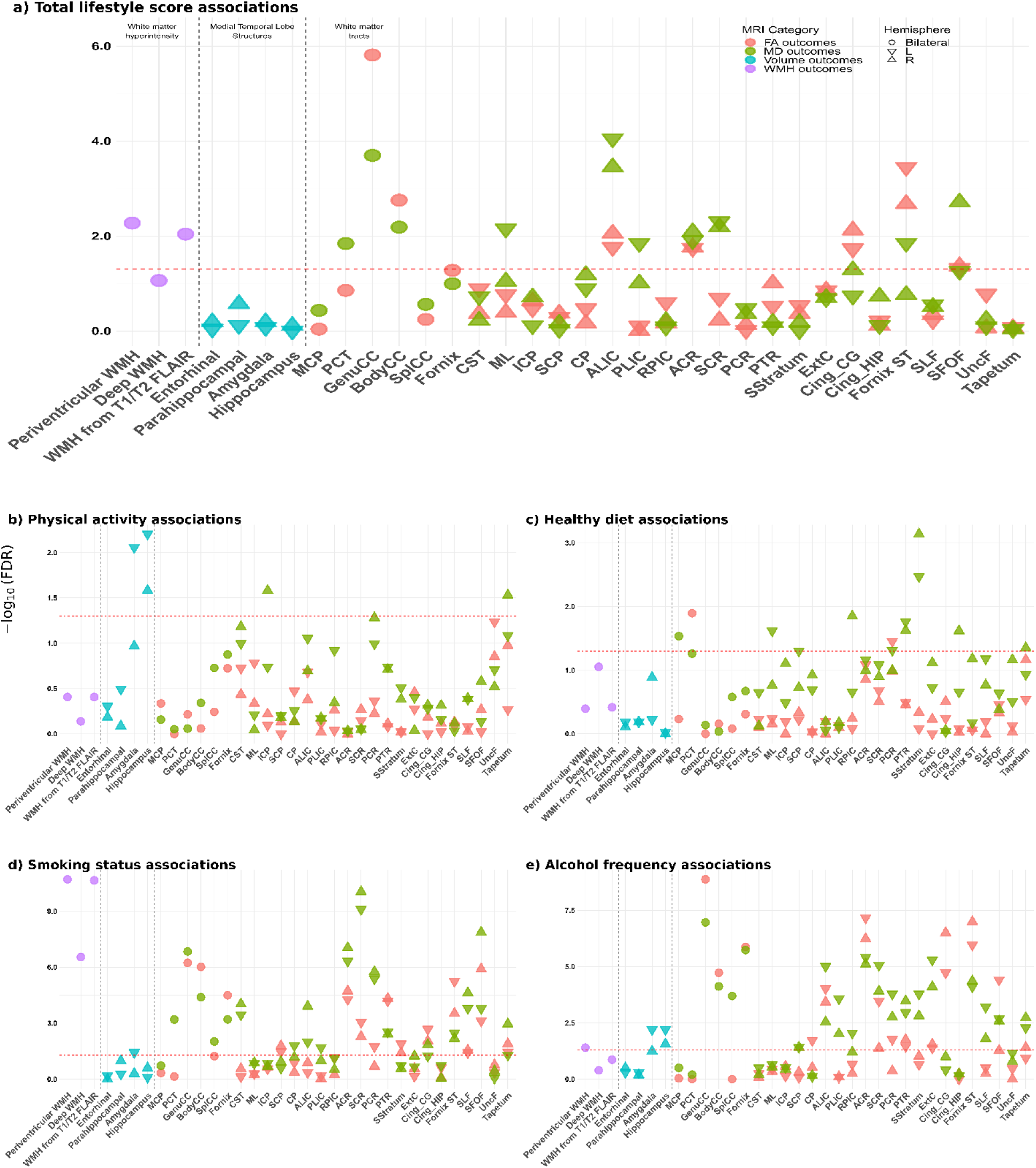
Manhattan plot of the associations between lifestyle factors (composite score and individual components) and MRI markers. The horizontal line indicates the threshold for multiple comparison correction (e.g., FDR-adjusted p < 0.05). ***Abbreviations*** *are listed at the end of the manuscript*.

### Interaction effects of APOE (ε2/ε4) dosages and healthy lifestyle on MRI

### outcomes

Prior to interaction analyses, no significant correlation was observed between *APOE* ε4/ε2 dosage and TLS (Spearman’s ρ = –0.006/–0.007; p = 0.369/0.304)

We examined the interaction between *APOE* dosages (ε2/ε4) and TLS as a continuous variable on neuroimaging outcomes. No FDR-significant interactions were observed (**Supplementary Table 12**). Similarly, when TLS was categorized into three groups (Poor, Intermediate, and Optimal; n = 5146, 9084, and 6784, respectively), no FDR-significant interaction effects were found (**Supplementary Table 13**).

In secondary analyses, no FDR-significant interactions were found between APOE dosages and individual binary lifestyle factors (**Supplementary Table 14**).

#### APOE Genotypes

No significant interaction effects were observed between APOE genotype and the TLS, assessed either as a continuous or categorical variable, or with any individual lifestyle component (**Supplementary Tables 15–16**).

### Sensitivity analyses

We conducted several sensitivity analyses to evaluate the robustness of our findings. First, repeating the models with a partially adjusted covariate set (excluding socioeconomic and health status variables) yielded results that were consistent with the main analyses, with no notable changes observed in the associations or interaction effects. Second, reclassifying the TLS into a binary variable (healthy: score = 3-4 vs. non-healthy: score = 0– 2) instead of using the original three-level classification produced similar results, with no significant *APOE* × lifestyle interactions detected (*p*_FDR_ > 0.05). Third, we assessed the interaction between *APOE* genotype group and the quantitative dietary score (ranging from 0 to 7), the only individual lifestyle component available as a continuous measure. No significant interaction effects were observed for dietary score on neuroimaging outcomes (*p*_FDR_ > 0.05). Overall, these sensitivity analyses support the robustness of our primary findings.

Overall, these sensitivity analyses support the robustness of our primary findings.

## Discussion

In this large-scale analysis of neuroimaging data from 21,014 UKB participants, we examined the associations of APOE (ε2/ε4 dosage) and lifestyle factors with neuroimaging biomarkers relevant to AD, including MTL volumes, WMH, and WM integrity (FA and MD). APOE ε4 dosage was significantly associated with lower FA and higher MD in critical cognitive tracts such as the the cingulum (hippocampus), SStratum, PTR, as well as smaller hippocampal and parahippocampal volumes. Healthier lifestyles, especially non-smoking and lower alcohol intake, correlated with reduced WMH burden and improved WM integrity. No significant APOE × lifestyle interactions were observed, supporting a more direct effect of APOE ε4 on neurodegeneration rather than an interaction-based vulnerability pathway.

### Association of *APOE (*both ε2/ε4 dosage and genotype-based) with neuroimaging outcomes

Our findings demonstrate a strong and statistically significant association between *APOE* ε4 dosage and multiple neuroimaging outcomes, including lower WM integrity, as reflected by lower FA and higher MD in the Cing_HIP, PTR and SStratum, consistent with previous research [12]. In particular, WM changes in the ventral cingulum may reflect early microstructural disruption, potentially involving reduced myelin integrity or fiber coherence. Given its role in connecting key memory-related regions, structural disruption in this tract has been associated with early neuronal loss and neurofibrillary tangle (NFT) pathology [42]. Although NFT pathology generally precedes significant WM degeneration in Alzheimer’s disease, the observed diffusion changes likely represent downstream effects rather than the primary cause of neurodegeneration. *APOE*-related changes in the PTR may reflect ventricular expansion, a hallmark of AD [43]. Given the PTR’s anatomical proximity to the lateral ventricles, observed WM alterations may reflect ventricular expansion and periventricular atrophy, processes commonly seen in AD. However, further studies are needed to confirm whether these microstructural changes in PTR are causally linked to APOE-related neurodegeneration and cognitive outcomes [12]. For SStratum, which is among the regions most susceptible to ε4-related axonal degeneration and demyelination. This vulnerability may stem from ε4-related disruptions in lipid metabolism and compromised myelin repair. As a major long-range association fiber pathway connecting the occipital, temporal, and frontal lobes, the SStratum supports a wide range of higher-order cognitive functions, including semantic processing, visual recognition, spatial navigation, and visual memory. Damage to this tract may therefore contribute to cognitive decline in ε4 carriers [18,44].

In addition, we observed significant *APOE* ε4-related effects in the fonix ST, PLIC, cingulum CC, MCP, SLF, corpus callosum, ALCI, SCR. Despite their anatomical diversity, these tracts share functional relevance to cognitive and emotional processing, and several are closely connected to the limbic system or subcortical-cortical circuits implicated in early AD, those WM region among the most affected regions in AD, showing top rank associations with memory impairment, amyloid-beta accumulation, and clinical cognitive decline [45]. Their shared vulnerability may reflect *APOE* ε4’s broader impact on neurodegenerative processes affecting long-range associative fibers, supporting its role in widespread WM disruption.

Additionally, we found that not only APOE ε4 dose but also the ε4/ε4 genotype was significantly associated with reduced bilateral hippocampal volume. We also observed associations with the right parahippocampal volume (dose model) and the bilateral amygdala volumes (genotype model). These findings are consistent with some previous studies, such as Breddels et al. [46], which reported APOE4-related differences in hippocampus and amygdala volumes. In contrast, Lyall et al. [14] did not find such associations in the UKB, possibly due to limited sample size (n ≈ 8,000) and differences in modeling. Our findings further highlight the sensitivity of the dosage model in detecting subtle structural changes, in line with histopathological evidence showing early neuronal loss in grey matter regions, especially within the medial temporal lobe, in AD [47]. Regarding the association between APOE and WMH, the genotype model revealed an FDR-significant effect of the ε4/ε4 genotype on deep WMH volume. Individuals homozygous for APOE ε4 may be particularly susceptible to vascular pathology. This aligns with previous research [12], further supporting the link between APOE genotype and WMH burden. Given that WMHs are widely recognized as markers of cerebral small vessel disease [48], our results contribute to the growing evidence that APOE may influence AD risk not only through amyloid and tau pathways, but also via vascular mechanisms.

### Association of lifestyle with neuroimaging outcomes

Beyond genetic influences, we observed strong associations between lifestyle factors and the widespread brain imaging metrics. A higher total healthy lifestyle score was linked to better WM integrity across multiple brain regions, including the corpus callosum, corticospinal tracts, and associative fibers such as the superior longitudinal fasciculus and fronto-occipital fasciculus. Among individual lifestyle components, non-smoking status and lower alcohol consumption had the most pronounced effects, contributing to reduced WMH burden and improved WM microstructure, in corpus callosum, corona radiata, fornix, and cingulum. These findings align with prior research demonstrating that smoking and excessive alcohol consumption are significant risk factors for WM deterioration and increased WMH burden [49,50]. Interestingly, a lower frequency of alcohol consumption was associated with larger hippocampal and amygdala volumes, suggesting a potential absence of alcohol-related neurotoxicity, preserving brain structures critical for memory and emotional regulation. This is consistent with evidence suggesting that alcohol accelerates hippocampal atrophy and neurodegeneration, likely through mechanisms such as oxidative stress, vascular dysfunction, and inflammation [51]. This finding reinforces public health recommendations to reduce alcohol intake for long-term brain health.

### G × E Interactions

In our study, no significant interactions between APOE and lifestyle factors were observed across any model specification—whether using APOE ε4 dosage or genotype models, and regardless of whether lifestyle was treated as a continuous score, categorical variable, or examined by individual components. This consistent null finding suggests that interventions targeting lifestyle improvements may be similarly beneficial for individuals irrespective of their APOE genotype, at least within this population. Our results support a similar pattern observed in previous large-scale studies investigating APOE-lifestyle interactions on cognition, where no significant interactions were reported [19]. Likewise, this lack of interaction has also been observed in studies on coronary artery disease [52], suggesting that the influence of lifestyle factors on health outcomes may not strongly depend on APOE genotype across different disease domains.

Nevertheless, a previous neuroimaging study with a relatively small sample size (n = 134) reported significant interactions between APOE ǫ4 status and lifestyle factors, such as diet and cognitive activity, on multimodal neuroimaging outcomes (p-values < .04) [53]. Some studies have also explored whether APOE ǫ4 moderates the effects of physical activity on brain imaging markers [54]. However, these findings may be limited by low statistical power and potentially influenced by publication bias. Given the larger sample size and greater statistical power of our study, our findings may provide a more reliable estimate of the absence of interaction effects. It should be noted, however, that certain gene– environment interactions may have gone undetected due to limitations such as the “healthy volunteer” selection bias commonly observed in the UKB cohort [55]. Additionally, a genome-wide association study identified APOE as one of the strongest genetic correlates of physical activity, particularly among older adults [56], suggesting that APOE ǫ4 carriers may be more health-conscious than the general population. Such genotype-related behavioral differences could further obscure potential gene–lifestyle interactions. Furthermore, our analysis focused on conventional lifestyle factors, and did not consider other domains (e.g., social engagement or mindfulness), where preliminary evidence for gene–environment interaction has been reported [57].

### Strengths and Limitations

The major strength of this study lies in utilizing data from the UKB, which, to our knowledge, represents the largest investigation to date exploring the associations between APOE dosage, lifestyle factors, and neuroimaging outcomes. Additionally, we provided a comprehensive examination of APOE genotype effects, complementing the primary dosage-based analyses. This extensive and well-characterized dataset enhances the robustness and reliability of our findings, allowing for precise estimation of effects across multiple genetic backgrounds.

Several limitations should be considered when interpreting our findings. First, although the UKB dataset provides a large and well-characterized sample, it is not fully representative of the general population, as participants tend to be healthier, more educated, and socioeconomically advantaged [55], which may limit the generalizability of our results. Second, the cross-sectional nature of this study precludes causal inferences; longitudinal analyses will be necessary to determine whether observed associations predict cognitive decline or AD progression over time. Additionally, although DTI metrics such as FA and MD provide valuable insights into WM integrity, they do not directly measure specific microstructural changes, such as axonal density or myelin integrity. Rather, they reflect the combined effects of multiple microstructural properties. For example, FA is influenced by axonal density, fiber coherence, and myelin integrity, while MD reflects changes in tissue density, extracellular space, and overall microstructural degradation. As emphasized by Jones et al. [42], diffusion metrics are sensitive but nonspecific indicators of microstructural changes and should be interpreted with caution [58]. The use of advanced neuroimaging techniques, such as neurite orientation dispersion and density imaging (NODDI) or myelin-sensitive MRI, could offer a more comprehensive understanding of WM alterations in relation to APOE and lifestyle factors.

Despite these limitations, our results carry important implications. We observed direct associations between APOE ε4 dosage and neuroimaging markers, and consistent benefits of healthy lifestyle behaviors across genotypes. The lack of significant interaction effects suggests that APOE and lifestyle contribute independently to brain structural integrity. This underscores the potential value of universally applicable lifestyle-based recommendations rather than genotype-specific advice for brain health.

## Conclusion

In conclusion, our study found robust associations between APOE ε4—examined using both dosage and genotype models—and multiple adverse brain MRI markers, including reduced medial temporal lobe volumes, altered WM integrity, and increased WMH. While healthier lifestyle factors were consistently associated with more favorable brain outcomes, we found no evidence that these factors mitigated the detrimental effects of APOE ε4. These findings suggest that APOE ε4 exerts an independent effect on brain structure, reinforcing its role as a key genetic risk factor. Nonetheless, our results highlight the universal benefits of healthy lifestyle behaviors for brain health, regardless of genetic background.

## Supporting information

Suppemental main text

## Data Availability

All data produced in the present study are available upon reasonable request to the authors

## Acknowledgments

This research has been conducted using the UKB Resource under Application Number 55392. Author Y. Zhang was supported by the China Scholarship Council (CSC) from the Ministry of Education of P.R. China. Authors S. Guloksuz and B.P.F. Rutten received support from the YOUTH-GEMs project, funded by the European Union’s Horizon Europe program under the grant agreement number: 101057182. S. Guloksuz was supported by the Ophelia research project, ZonMw grant number: 636340001. D. van der Meer is supported by the Research Council of Norway (RCN #351751). We declare that there are no conflicts of interest among the authors.

## Abbreviations Used in Figures

Abbreviations used in the figures are as follows:

GenuCC: Genu of Corpus Callosum
BodyCC: Body of Corpus Callosum
SplCC: Splenium of Corpus Callosum
MCP: Middle Cerebellar Peduncle
ICP: Inferior Cerebellar Peduncle
SCP: Superior Cerebellar Peduncle
PCT: Pontine Crossing Tract
CP: Cerebral Peduncle
ALIC: Anterior Limb of Internal Capsule
PLIC: Posterior Limb of Internal Capsule
RPIC: Retrolenticular Part of Internal Capsule
ACR: Anterior Corona Radiata
SCR: Superior Corona Radiata
PCR: Posterior Corona Radiata
CST: Corticospinal Tract
ML: Medial Lemniscus
SLF: Superior Longitudinal Fasciculus
SFOF: Superior Fronto-Occipital Fasciculus
UncF: Uncinate Fasciculus
Cing_CG: Cingulum Cingulate Gyrus
Cing_HIP: Cingulum Hippocampus
Fornix: Fornix
Fornix ST: fornix/stria terminalis
PTR: Posterior Thalamic Radiation
SStratum: Sagittal Stratum
ExtC: External Capsule.

## Notes

### Competing Interest Statement

The authors have declared no competing interest.

### Funding Statement

Funding Statement:
Author Y. Zhang was supported by the China Scholarship Council (CSC) from the Ministry of Education of the People's Republic of China. Authors S. Guloksuz and B.P.F. Rutten received support from the YOUTH-GEMs project, funded by the European Union's Horizon Europe program (grant agreement number: 101057182). S. Guloksuz was also supported by the Ophelia research project (ZonMw grant number: 636340001). D. van der Meer is supported by the Research Council of Norway (grant number: 351751).
Conflict of Interest:
The authors declare that there are no conflicts of interest.

### Author Declarations

Ethics committee/IRB of the North West Multi-centre Research Ethics Committee (REC reference 11/NW/0382) gave ethical approval for this work. All participants in the UK Biobank study provided electronic informed consent. Data access for this study was obtained under UK Biobank application number 55392.

## References

[1] Alzheimer’s Association et al. “2012 Alzheimer’s disease facts and figures”. In: Alzheimer’s & dementia 8.2 (2012), pp. 131–168.

[2] Amanda M Di Battista, Nicolette M Heinsinger, and G William Rebeck. “Alzheimer’s disease genetic risk factor APOE-ε4 also affects normal brain function”. In: Current Alzheimer Research 13.11 (2016), pp. 1200–1207.

[3] Benjamin R Troutwine, et al. “Apolipoprotein E and Alzheimer’s disease”. In: Acta Pharmaceutica Sinica B 12.2 (2022), pp. 496–510.

[4] David M Holtzman, Joachim Herz, and Guojun Bu. “Apolipoprotein E and apolipoprotein E receptors: normal biology and roles in Alzheimer disease”. In: Cold Spring Harbor perspectives in medicine 2.3 (2012), a006312.

[5] John C Morris. “Early-stage and preclinical Alzheimer disease.” In: Alzheimer disease and associated disorders 19.3 (2005), pp. 163–165.

[6] Freddie Márquez and Michael A Yassa. “Neuroimaging biomarkers for Alzheimer’s disease”. In: Molecular neurodegeneration 14.1 (2019), p. 21.

[7] Latha Velayudhan et al. “Entorhinal cortex thickness predicts cognitive decline in Alzheimer’s disease”. In: Journal of Alzheimer’s Disease 33.3 (2013), pp. 755–766.

[8] Brian T Gold, et al. “White matter integrity and vulnerability to Alzheimer’s disease: preliminary findings and future directions”. In: Biochimica et Biophysica Acta (BBA)-Molecular Basis of Disease 1822.3 (2012),pp. 416–422.

[9] David Medina, et al. “White matter changes in mild cognitive impairment and AD: A diffusion tensor imaging study”. In: Neurobiology of aging 27.5 (2006), pp. 663–672.

[10] Talia M Nir, et al. “Effectiveness of regional DTI measures in distinguishing Alzheimer’s disease, MCI, and normal aging”. In: NeuroImage: clinical 3 (2013), pp. 180–195.

[11] Michele Veldsman, et al. “The human hippocampus and its subfield volumes across age, sex and APOE e4 status”. In: Brain Communications 3.1 (2021), fcaa219.

[12] Verena Heise, et al. “A comprehensive analysis of APOE genotype effects on human brain structure in the UK Biobank”. In: Translational Psychiatry 14.1 (2024), p. 143.

[13] Donald M Lyall, et al. “Alzheimer’s disease susceptibility genes APOE and TOMM40, and hippocampal volumes in the Lothian birth cohort 1936”. In: PloS one 8.11 (2013), e80513.

[14] Donald M Lyall, et al. “Association between APOE e4 and white matter hyperintensity volume, but not total brain volume or white matter integrity”. In: Brain imaging and behavior 14 (2020), pp. 1468–1476.

[15] Jinping Sun, et al. “APOE ε4 allele accelerates age-related multi-cognitive decline and white matter damage in non-demented elderly”. In: Aging (Albany NY) 12.12 (2020), p. 12019.

[16] Yuesong Pan et al. “Adherence to a healthy lifestyle and brain structural imaging markers”. In: European journal of epidemiology 38.6 (2023), pp. 657–668.

[17] Nora Bittner, et al. “When your brain looks older than expected: combined lifestyle risk and BrainAGE”. In: Brain Structure and Function 226 (2021), pp. 621–645.

[18] Chia-Chen Liu, et al. “Apolipoprotein E and Alzheimer disease: risk, mechanisms and therapy”. In: Nature Reviews Neurology 9.2 (2013), pp. 106–118.

[19] Donald M Lyall, et al. “Assessing for interaction between APOE ε4, sex, and lifestyle on cognitive abilities”. In: Neurology 92.23 (2019), e2691–e2698.

[20] J Carson Smith, et al. “Interactive effects of physical activity and APOE-ε4 on white matter tract diffusivity in healthy elders”. In: Neuroimage 131 (2016), pp. 102–112.

[21] J Carson Smith, et al. “Physical activity reduces hippocampal atrophy in elders at genetic risk for Alzheimer’s disease”. In: Frontiers in aging neuroscience 6 (2014), p. 61.

[22] Cathie Sudlow, et al. “UK biobank: an open access resource for identifying the causes of a wide range of complex diseases of middle and old age”. In: PLoS medicine 12.3 (2015), e1001779.

[23] Thomas J Littlejohns, et al. “The UK Biobank imaging enhancement of 100,000 participants: rationale, data collection, management and future directions”. In: Nature communications 11.1 (2020), p. 2624.

[24] Clare Bycroft, et al. “The UK Biobank resource with deep phenotyping and genomic data”. In: Nature 562.7726 (2018), pp. 203–209.

[25] Marius De Groot, et al. “Improving alignment in tract-based spatial statistics: evaluation and optimization of image registration”. In: Neuroimage 76 (2013), pp. 400–411.

[26] Shreeya S Navale et al. “Vitamin D and brain health: an observational and Mendelian randomization study”. In: The American journal of clinical nutrition 116.2 (2022), pp. 531– 540.

[27] May A Beydoun, et al. “Cardiovascular health, infection burden and their interactive association with brain volumetric and white matter integrity outcomes in the UK Biobank”. In: Brain, Behavior, and Immunity 113 (2023), pp. 91–103.

[28] Ludovica Griffanti, et al. “BIANCA (Brain Intensity AbNormality Classification Algorithm): A new tool for automated segmentation of white matter hyperintensities”. In: Neuroimage 141 (2016), pp. 191–205.

[29] Karla L Miller, et al. “Multimodal population brain imaging in the UK Biobank prospective epidemiological study”. In: Nature neuroscience 19.11 (2016), pp. 1523–1536.

[30] Ludovica Griffanti, et al. “Classification and characterization of periventricular and deep white matter hyperintensities on MRI: a study in older adults”. In: Neuroimage 170 (2018), pp. 174–181.

[31] Fidel Alfaro-Almagro, et al. “Image processing and Quality Control for the first 10,000 brain imaging datasets from UK Biobank”. In: Neuroimage 166 (2018), pp. 400–424.

[32] Anwar Mulugeta, et al. “Healthy lifestyle, genetic risk and brain health: A gene-environment interaction study in the UK biobank”. In: Nutrients 14.19 (2022), p. 3907.

[33] Emelia J Benjamin et al. “Heart disease and stroke statistics—2017 update: a report from the American Heart Association”. In: circulation 135.10 (2017), e146–e603.

[34] Ilianna Lourida, et al. “Association of lifestyle and genetic risk with incidence of dementia”. In: Jama 322.5 (2019), pp. 430–437.

[35] Piril Hepsomali and John A Groeger. “Diet and general cognitive ability in the UK Biobank dataset”. In: Scientific reports 11.1 (2021), p. 11786.

[36] Dariush Mozaffarian. “Dietary and policy priorities for cardiovascular disease, diabetes, and obesity: a comprehensive review”. In: Circulation 133.2 (2016), pp. 187–225.

[37] Claire T McEvoy et al. “Neuroprotective diets are associated with better cognitive function: the health and retirement study”. In: Journal of the American Geriatrics Society 65.8 (2017), pp. 1857–1862.

[38] Julian Mutz, Charlotte J Roscoe, and Cathryn M Lewis. “Exploring health in the UK Biobank: associations with sociodemographic characteristics, psychosocial factors, lifestyle and environmental exposures”. In: BMC medicine 19 (2021), pp. 1–18.

[39] Franz Faul, et al. “G* Power 3: A flexible statistical power analysis program for the social, behavioral, and biomedical sciences”. In: Behavior research methods 39.2 (2007), pp. 175–191.

[40] Nathan Pike. “Using false discovery rates for multiple comparisons in ecology and evolution”. In: Methods in ecology and Evolution 2.3 (2011), pp. 278–282.

[41] Simon R Cox, et al. “Structural brain imaging correlates of general intelligence in UK Biobank”. In: Intelligence 76 (2019), p. 101376.

[42] Jinyu Zhou, et al. “White matter damage in Alzheimer’s disease: contribution of oligodendrocytes”. In: Current Alzheimer Research 19.9 (2022), pp. 629–640.

[43] Jean-Philippe Coutu et al. “White matter changes are associated with ventricular expansion in aging, mild cognitive impairment, and Alzheimer’s disease”. In: Journal of Alzheimer’s Disease 49.2 (2015), pp. 329–342.

[44] Ahmed Adnan Al-Juboori, et al. “Clinical implications of sagittal stratum damage: Laterality, neuroanatomical developmental considerations, and functional outcomes”. In: Surgical Neurology International 16 (2025), p. 4.

[45] Emrin Horgusluoglu-Moloch, et al. “Systems modeling of white matter microstructural abnormalities in Alzheimer’s disease”. In: NeuroImage: Clinical 26 (2020), p. 102203.

[46] Esmee M Breddels et al. “Brain morphology mediating the effects of common genetic risk variants on Alzheimer’s disease”. In: Journal of Alzheimer’s Disease Reports 9 (2025), p. 25424823251328300.

[47] Alberto Serrano-Pozo, et al. “Neuropathological alterations in Alzheimer disease”. In: Cold Spring Harbor perspectives in medicine 1.1 (2011), a006189.

[48] Annemieke Ter Telgte, et al. “Cerebral small vessel disease: from a focal to a global perspective”. In: Nature Reviews Neurology 14.7 (2018), pp. 387–398.

[49] Keshuo Lin et al. “Risk factors and cognitive correlates of white matter hyperintensities in ethnically diverse populations without dementia: The COSMIC consortium”. In: Alzheimer’s & Dementia: Diagnosis, Assessment & Disease Monitoring 16.1 (2024), e12567.

[50] Vera Thornton, et al. “Alcohol, smoking, and brain structure: common or substance specific associations”. In: medRxiv (2024).

[51] Anya Topiwala, et al. “Alcohol consumption and MRI markers of brain structure and function: cohort study of 25,378 UK Biobank participants”. In: NeuroImage: Clinical 35 (2022), p. 103066.

[52] Maxime M Bos, et al. “Apolipoprotein E genotype, lifestyle and coronary artery disease: Gene-environment interaction analyses in the UK Biobank population”. In: Atherosclerosis 328 (2021), pp. 33–37.

[53] Francesca Felisatti, et al. “Interaction between APOE4 and lifestyle on neuroimaging biomarkers and cognition in cognitively unimpaired older adults”. In: Alzheimer’s & Dementia 19 (2023), e062118.

[54] Jaisalmer de Frutos-Lucas, et al. “Does APOE genotype moderate the relationship between physical activity, brain health and dementia risk? A systematic review”. In: Ageing Research Reviews 64 (2020), p. 101173.

[55] Anna Fry et al. “Comparison of sociodemographic and health-related characteristics of UK Biobank participants with those of the general population”. In: American journal of epidemiology 186.9 (2017), pp. 1026– 1034.

[56] Yann C Klimentidis et al. “Genome-wide association study of habitual physical activity in over 377,000 UK Biobank participants identifies multiple variants including CADM2 and APOE”. In: International journal of obesity 42.6 (2018), pp. 1161–1176.

[57] Deirdre M O’Shea, et al. “APOE ε4 carrier status moderates the effect of lifestyle factors on cognitive reserve”. In: Alzheimer’s & dementia 20.11 (2024), pp. 8062–8073.

[58] Derek K Jones, Thomas R Knösche, and Robert Turner. “White matter integrity, fiber count, and other fallacies: the do’s and don’ts of diffusion MRI”. In: Neuroimage 73 (2013), pp. 239–254.

