## Supplementary material for "Associations of *APOE* e2/e4 dosage and lifestyle on neuroimaging markers in the UK Biobank": Suppemental main text

### Supplementary Tables

#### Supplementary Table 1. Selected Neuroimaging Measures: sMRI and dMRI Phenotypes

(See Supplementary Excel File, Sheet 1)

#### Supplementary Table 2. Descriptions of lifestyle factors included in the analyses.

| **Lifestyle factors** | **UK Biobank field ID** | **Categorisation or description** | **Remarks** |
| --- | --- | --- | --- |
| Alcohol frequency | 1558 ["About how often do you drink alcohol?"] | Non-drinker  Special occasions  1-3 times/month  1 or 2 times/week  3 0r 4 times/week  Daily or almost daily | **NOT** “Daily or almost daily" was coded as 1. |
| Smoking | 20116 [Current/past smoking status] | Non-smokers  Ex-smokers  Current smokers | **NOT** a "Current smoker" was coded as 1 |
| Regular physical activity | 884 [In a typical WEEK, on how many days did you do 10 minutes  or more of moderate physical activities like carrying light loads,  cycling at a normal pace? (Do not include walking)]  894 [How many minutes did you usually spend doing moderate  activities on a typical DAY?] | Yes (1)  No (0) | individuals who met either of the below  criteria were coded “Yes” otherwise “No”  for the regular physical activity.  - Moderate physical activity at least for  150 minute per week  OR  - Vigorous physical activity for at least  75 minutes per week |
| Diet |  | healthy diet (1)  unhealthy diet (0) | Total score > 4 classified as a healthy diet (coded as 1), score ≤ 4 coded as 0 |

#### Supplementary Table 3. Descriptions of diet score in the analyses.

| **Diet component** | **Intake goal** | | **Field IDs** | **Amount per serving** |
| --- | --- | --- | --- | --- |
| Fruit | >=3 serving/day | 1309 (pieces fresh fruit/day) 1319 (pieces dried fruit/day) | | 1309 – 1 piece 1319 – 5 pieces |
| Vegetable | >=3 servings/day | 1289 (tablespoons cooked vegetables/day) 1299 (salad/raw vegetables/day) | | 3 tablespoons-1serving |
| Whole grains | >=3 servings/day | 1438, 1448 (wholemeal/wholegrain bread  slices/week)  1458, 1468 (bran/oat/muesli cereal bowls/week) | | 1438/1448 – 1 slice/day  1458/1468 – 1 bowl/day |
| (Shell)fish | ≥2  servings/week | 1329 (oily fish/week)  1339 (non-oily fish/week) | |  |
| Refined grains | ≤1.5 servings/day | 1438, 1448 (white, brown, other bread slices/week)  1458, 1468 (biscuit, other cereals/week) | | 1438/1448 – 1 slice/day  1458/1468 – 1 bowl/day |
| Processed meats | ≤2 serving/week | 1349 (processed meat/week or daily) | | less than twice a week |
| Unprocessed meats | ≤ 1.5 serving/wk | 1369 (beef/week or day) 1379 (lamb or mutton/week or day) 1389 (pork/week or day) | | “never”=0, “less than once a week”=0.5,  “once a week”=1, “2-4 times a week”=3, “5-6 times a week”=5.5,  “once or more daily”=7 |

#### Supplementary Table 4. Excluded (self-reported) diseases.

| **Supplementary Table 1** Excluded (self-reported) diseases. | coding |
| --- | --- |
| Brain haemorrhage | 1491 |
| brain abscess/intracranial abscess | 1245 |
| Cerebral aneurysm | 1425 |
| Cerebral palsy | 1433 |
| Chronic/degenerative neurological problem | 1258 |
| Dementia/Alzheimer disease/cognitive impairment | 1263 |
| Encephalitis | 1246 |
| Epilepsy | 1264 |
| Fracture skull/head | 1626 |
| Head injury | 1266 |
| Infection of nervous system | 1244 |
| Ischaemic stroke | 1583 |
| meningioma / benign meningeal tumour | 1659 |
| Meningitis | 1247 |
| Motor neurone disease | 1259 |
| Multiple sclerosis | 1261 |
| Neurological injury/trauma | 1240 |
| benign neuroma | 1683 |
| Other demyelinating condition | 1397 |
| Other neurological problem | 1434 |
| Parkinson disease | 1262 |
| Spina bifida | 1524 |
| Stroke | 1081 |
| Subarachnoid haemorrhage | 1086 |
| Subdural haematoma | 1083 |
| Transient ischaemic attack | 1082 |

#### Supplementary Table 5. Characteristics of the study sample and comparison of participants that were included and excluded

| **Characteristic** | **Include  N = 21,014^1^** | **Exclude  N = 481,342^1^** | **p-value^2^** |
| --- | --- | --- | --- |
| Age | 54.596 (7.488) | 56.614 (8.110) | <0.001 |
| Sex |  |  | <0.001 |
| Female | 10,507 (50%) | 262,787 (55%) |  |
| Male | 10,507 (50%) | 218,555 (45%) |  |
| Education |  |  | <0.001 |
| College/University Degree | 10,214 (49%) | 152,305 (32%) |  |
| Education to age 18 or above | 4,833 (23%) | 109,609 (23%) |  |
| Education to age 16 qualifications | 4,833 (23%) | 128,151 (27%) |  |
| No qualifications | 1,134 (5.4%) | 84,885 (18%) |  |
| Unknown | 0 | 6,392 |  |
| Income Status |  |  | <0.001 |
| < £18,000 | 2,220 (11%) | 94,954 (23%) |  |
| £18,000–£30,999 | 4,498 (21%) | 103,639 (26%) |  |
| £31,000–£51,999 | 6,322 (30%) | 104,423 (26%) |  |
| £52,000–£100,000 | 6,210 (30%) | 80,030 (20%) |  |
| > £100,000 | 1,764 (8.4%) | 21,159 (5.2%) |  |
| Unknown | 0 | 77,137 |  |
| Employment Status |  |  | <0.001 |
| Employed | 14,854 (71%) | 272,187 (57%) |  |
| Retired | 5,152 (25%) | 161,809 (34%) |  |
| Unemployed | 1,008 (4.8%) | 41,596 (8.7%) |  |
| Unknown | 0 | 5,750 |  |
| Longstanding Illness |  |  | <0.001 |
| No | 16,691 (79%) | 312,460 (67%) |  |
| Yes | 4,323 (21%) | 155,526 (33%) |  |
| Unknown | 0 | 13,356 |  |
| Alcohol Frequency |  |  | <0.001 |
| High Frequency | 4,825 (23%) | 96,919 (20%) |  |
| Low Frequency | 16,189 (77%) | 382,921 (80%) |  |
| Unknown | 0 | 1,502 |  |
| Smoking Status |  |  | <0.001 |
| Current smoker | 1,145 (5.4%) | 51,813 (11%) |  |
| Non-smoker | 19,869 (95%) | 426,579 (89%) |  |
| Unknown | 0 | 2,950 |  |
| Healthy Diet |  |  | <0.001 |
| Unhealthy | 9,888 (47%) | 262,931 (55%) |  |
| Healthy | 11,126 (53%) | 218,411 (45%) |  |
| Physical Activity |  |  | <0.001 |
| No | 4,425 (21%) | 50,503 (18%) |  |
| Yes | 16,589 (79%) | 236,613 (82%) |  |
| Unknown | 0 | 194,226 |  |
| Total Healthy Lifestyle Score |  |  | 0.004 |
| 0 | 50 (0.2%) | 880 (0.3%) |  |
| 1 | 807 (3.8%) | 11,949 (4.2%) |  |
| 2 | 4,289 (20%) | 59,352 (21%) |  |
| 3 | 9,084 (43%) | 124,213 (43%) |  |
| 4 | 6,784 (32%) | 89,703 (31%) |  |
| Unknown | 0 | 195,245 |  |
| ^1^Mean (SD); n (%) | | | |
| ^2^Wilcoxon rank sum test; Pearson's Chi-squared test with simulated p-value (based on 2000 replicates) | | | |

#### Supplementary Table 6. Baseline Characteristics of Participants by APOE Genotype (The FA and MD metrics)

| **Characteristic** | **Overall  N = 21,014^1^** | **e2e2  N = 120^1^** | **e2e3  N = 2,638^1^** | **e2e4  N = 499^1^** | **e3e3  N = 12,355^1^** | **e3e4  N = 4,932^1^** | **e4e4  N = 470^1^** | **p-value^2^** |
| --- | --- | --- | --- | --- | --- | --- | --- | --- |
| Middle Cerebellar Peduncle FA | 0.5450 (0.0213) | 0.5478 (0.0199) | 0.5455 (0.0214) | 0.5455 (0.0209) | 0.5452 (0.0213) | 0.5444 (0.0214) | 0.5429 (0.0210) | 0.028 |
| Pontine Crossing Tract FA | 0.4061 (0.0321) | 0.4074 (0.0307) | 0.4062 (0.0320) | 0.4059 (0.0312) | 0.4062 (0.0320) | 0.4057 (0.0325) | 0.4052 (0.0320) | 0.937 |
| Genu of Corpus Callosum FA | 0.7246 (0.0315) | 0.7249 (0.0317) | 0.7246 (0.0318) | 0.7257 (0.0310) | 0.7245 (0.0317) | 0.7248 (0.0311) | 0.7232 (0.0299) | 0.820 |
| Body of Corpus Callosum FA | 0.7154 (0.0266) | 0.7131 (0.0254) | 0.7160 (0.0266) | 0.7167 (0.0261) | 0.7154 (0.0267) | 0.7153 (0.0265) | 0.7141 (0.0264) | 0.313 |
| Splenium of Corpus Callosum FA | 0.7902 (0.0199) | 0.7894 (0.0186) | 0.7902 (0.0196) | 0.7918 (0.0204) | 0.7903 (0.0199) | 0.7901 (0.0198) | 0.7881 (0.0208) | 0.144 |
| Fornix FA | 0.4356 (0.0839) | 0.4449 (0.0744) | 0.4370 (0.0833) | 0.4373 (0.0829) | 0.4349 (0.0834) | 0.4360 (0.0857) | 0.4403 (0.0811) | 0.325 |
| Corticospinal Tract FA | 0.5308 (0.0320) | 0.5335 (0.0311) | 0.5308 (0.0320) | 0.5301 (0.0310) | 0.5309 (0.0318) | 0.5308 (0.0324) | 0.5279 (0.0324) | 0.513 |
| Medial Lemniscus FA | 0.5956 (0.0250) | 0.5929 (0.0254) | 0.5963 (0.0246) | 0.5956 (0.0242) | 0.5953 (0.0252) | 0.5961 (0.0247) | 0.5938 (0.0245) | 0.128 |
| Inferior Cerebellar Peduncle FA | 0.5516 (0.0262) | 0.5509 (0.0250) | 0.5524 (0.0261) | 0.5519 (0.0277) | 0.5513 (0.0263) | 0.5519 (0.0262) | 0.5524 (0.0243) | 0.306 |
| Superior Cerebellar Peduncle FA | 0.7059 (0.0223) | 0.7054 (0.0225) | 0.7060 (0.0222) | 0.7062 (0.0208) | 0.7060 (0.0225) | 0.7059 (0.0222) | 0.7045 (0.0223) | 0.769 |
| Cerebral Peduncle FA | 0.7119 (0.0211) | 0.7104 (0.0212) | 0.7121 (0.0210) | 0.7136 (0.0207) | 0.7118 (0.0213) | 0.7119 (0.0207) | 0.7103 (0.0202) | 0.252 |
| Anterior Limb of Internal Capsule FA | 0.5976 (0.0225) | 0.5968 (0.0245) | 0.5978 (0.0230) | 0.5989 (0.0215) | 0.5976 (0.0226) | 0.5974 (0.0222) | 0.5961 (0.0219) | 0.616 |
| Posterior Limb of Internal Capsule FA | 0.6847 (0.0224) | 0.6843 (0.0220) | 0.6842 (0.0225) | 0.6852 (0.0214) | 0.6848 (0.0225) | 0.6847 (0.0221) | 0.6842 (0.0218) | 0.837 |
| Retrolenticular Part of Internal Capsule FA | 0.6056 (0.0242) | 0.6069 (0.0246) | 0.6061 (0.0242) | 0.6064 (0.0241) | 0.6057 (0.0241) | 0.6053 (0.0243) | 0.6042 (0.0252) | 0.393 |
| Anterior Corona Radiata FA | 0.4585 (0.0285) | 0.4613 (0.0284) | 0.4589 (0.0291) | 0.4597 (0.0272) | 0.4583 (0.0286) | 0.4585 (0.0278) | 0.4586 (0.0288) | 0.695 |
| Superior Corona Radiata FA | 0.4873 (0.0231) | 0.4871 (0.0216) | 0.4874 (0.0236) | 0.4873 (0.0227) | 0.4872 (0.0231) | 0.4877 (0.0228) | 0.4881 (0.0238) | 0.834 |
| Posterior Corona Radiata FA | 0.4871 (0.0235) | 0.4875 (0.0241) | 0.4871 (0.0232) | 0.4880 (0.0231) | 0.4871 (0.0236) | 0.4870 (0.0235) | 0.4868 (0.0241) | 0.939 |
| Posterior Thalamic Radiation FA | 0.6006 (0.0305) | 0.6010 (0.0303) | 0.6015 (0.0306) | 0.6016 (0.0308) | 0.6006 (0.0304) | 0.6005 (0.0308) | 0.5967 (0.0303) | 0.057 |
| Sagittal Stratum FA | 0.5652 (0.0276) | 0.5665 (0.0296) | 0.5656 (0.0282) | 0.5673 (0.0267) | 0.5652 (0.0275) | 0.5653 (0.0275) | 0.5610 (0.0289) | 0.024 |
| External Capsule FA | 0.4637 (0.0216) | 0.4647 (0.0228) | 0.4635 (0.0219) | 0.4658 (0.0221) | 0.4636 (0.0216) | 0.4639 (0.0216) | 0.4630 (0.0208) | 0.297 |
| Cingulum Cingulate Gyrus FA | 0.6035 (0.0300) | 0.6029 (0.0304) | 0.6040 (0.0305) | 0.6032 (0.0304) | 0.6034 (0.0300) | 0.6034 (0.0299) | 0.6027 (0.0304) | 0.896 |
| Cingulum Hippocampus FA | 0.4624 (0.0320) | 0.4602 (0.0306) | 0.4630 (0.0318) | 0.4606 (0.0312) | 0.4628 (0.0320) | 0.4617 (0.0322) | 0.4563 (0.0319) | <0.001 |
| Fornix Cresstria Terminalis FA | 0.5255 (0.0322) | 0.5292 (0.0314) | 0.5258 (0.0320) | 0.5241 (0.0332) | 0.5255 (0.0321) | 0.5254 (0.0322) | 0.5245 (0.0335) | 0.905 |
| Superior Longitudinal Fasciculus FA | 0.5265 (0.0240) | 0.5283 (0.0240) | 0.5270 (0.0241) | 0.5266 (0.0241) | 0.5264 (0.0240) | 0.5264 (0.0241) | 0.5267 (0.0236) | 0.720 |
| Superior Frontooccipital Fasciculus FA | 0.4636 (0.0386) | 0.4636 (0.0382) | 0.4646 (0.0387) | 0.4634 (0.0392) | 0.4632 (0.0385) | 0.4640 (0.0387) | 0.4622 (0.0381) | 0.604 |
| Uncinate Fasciculus FA | 0.5199 (0.0336) | 0.5238 (0.0309) | 0.5202 (0.0341) | 0.5227 (0.0335) | 0.5196 (0.0338) | 0.5203 (0.0330) | 0.5183 (0.0329) | 0.145 |
| Tapetum FA | 0.5663 (0.0582) | 0.5670 (0.0597) | 0.5674 (0.0571) | 0.5705 (0.0615) | 0.5657 (0.0582) | 0.5666 (0.0583) | 0.5683 (0.0578) | 0.254 |
| Middle Cerebellar Peduncle MD | 0.0007 (0.0000) | 0.0007 (0.0000) | 0.0007 (0.0000) | 0.0007 (0.0000) | 0.0007 (0.0000) | 0.0007 (0.0000) | 0.0007 (0.0000) | 0.048 |
| Pontine Crossing Tract MD | 0.0008 (0.0001) | 0.0008 (0.0001) | 0.0008 (0.0001) | 0.0008 (0.0001) | 0.0008 (0.0001) | 0.0008 (0.0001) | 0.0008 (0.0001) | 0.165 |
| Genu of Corpus Callosum MD | 0.0008 (0.0000) | 0.0008 (0.0000) | 0.0008 (0.0000) | 0.0008 (0.0000) | 0.0008 (0.0000) | 0.0008 (0.0000) | 0.0008 (0.0000) | 0.515 |
| Body of Corpus Callosum MD | 0.0008 (0.0000) | 0.0008 (0.0000) | 0.0008 (0.0000) | 0.0008 (0.0000) | 0.0008 (0.0000) | 0.0008 (0.0000) | 0.0008 (0.0000) | 0.186 |
| Splenium of Corpus Callosum MD | 0.0007 (0.0000) | 0.0007 (0.0000) | 0.0007 (0.0000) | 0.0007 (0.0000) | 0.0007 (0.0000) | 0.0007 (0.0000) | 0.0007 (0.0000) | 0.225 |
| Fornix MD | 0.0016 (0.0003) | 0.0015 (0.0002) | 0.0016 (0.0003) | 0.0016 (0.0003) | 0.0016 (0.0003) | 0.0016 (0.0003) | 0.0016 (0.0003) | 0.240 |
| Corticospinal Tract MD | 0.0008 (0.0001) | 0.0008 (0.0001) | 0.0008 (0.0001) | 0.0008 (0.0001) | 0.0008 (0.0001) | 0.0008 (0.0001) | 0.0008 (0.0001) | 0.123 |
| Medial Lemniscus MD | 0.0008 (0.0000) | 0.0008 (0.0000) | 0.0008 (0.0000) | 0.0008 (0.0000) | 0.0008 (0.0000) | 0.0008 (0.0000) | 0.0008 (0.0000) | 0.010 |
| Inferior Cerebellar Peduncle MD | 0.0007 (0.0000) | 0.0007 (0.0000) | 0.0007 (0.0000) | 0.0007 (0.0000) | 0.0007 (0.0000) | 0.0007 (0.0000) | 0.0007 (0.0000) | 0.432 |
| Superior Cerebellar Peduncle MD | 0.0008 (0.0000) | 0.0008 (0.0000) | 0.0008 (0.0000) | 0.0008 (0.0000) | 0.0008 (0.0000) | 0.0008 (0.0000) | 0.0008 (0.0000) | 0.561 |
| Cerebral Peduncle MD | 0.0007 (0.0000) | 0.0007 (0.0000) | 0.0007 (0.0000) | 0.0007 (0.0000) | 0.0007 (0.0000) | 0.0007 (0.0000) | 0.0007 (0.0000) | 0.760 |
| Anterior Limb of Internal Capsule MD | 0.0007 (0.0000) | 0.0007 (0.0000) | 0.0007 (0.0000) | 0.0007 (0.0000) | 0.0007 (0.0000) | 0.0007 (0.0000) | 0.0007 (0.0000) | 0.379 |
| Posterior Limb of Internal Capsule MD | 0.0007 (0.0000) | 0.0007 (0.0000) | 0.0007 (0.0000) | 0.0007 (0.0000) | 0.0007 (0.0000) | 0.0007 (0.0000) | 0.0007 (0.0000) | 0.120 |
| Retrolenticular Part of Internal Capsule MD | 0.0008 (0.0000) | 0.0008 (0.0000) | 0.0008 (0.0000) | 0.0008 (0.0000) | 0.0008 (0.0000) | 0.0008 (0.0000) | 0.0008 (0.0000) | 0.045 |
| Anterior Corona Radiata MD | 0.0008 (0.0000) | 0.0008 (0.0000) | 0.0008 (0.0000) | 0.0008 (0.0000) | 0.0008 (0.0000) | 0.0008 (0.0000) | 0.0008 (0.0000) | 0.158 |
| Superior Corona Radiata MD | 0.0007 (0.0000) | 0.0007 (0.0000) | 0.0007 (0.0000) | 0.0007 (0.0000) | 0.0007 (0.0000) | 0.0007 (0.0000) | 0.0007 (0.0000) | 0.089 |
| Posterior Corona Radiata MD | 0.0008 (0.0000) | 0.0008 (0.0000) | 0.0008 (0.0000) | 0.0008 (0.0000) | 0.0008 (0.0000) | 0.0008 (0.0000) | 0.0008 (0.0000) | 0.099 |
| Posterior Thalamic Radiation MD | 0.0008 (0.0000) | 0.0008 (0.0000) | 0.0008 (0.0000) | 0.0008 (0.0000) | 0.0008 (0.0000) | 0.0008 (0.0000) | 0.0008 (0.0000) | 0.019 |
| Sagittal Stratum MD | 0.0008 (0.0000) | 0.0008 (0.0000) | 0.0008 (0.0000) | 0.0008 (0.0000) | 0.0008 (0.0000) | 0.0008 (0.0000) | 0.0008 (0.0000) | 0.001 |
| External Capsule MD | 0.0008 (0.0000) | 0.0008 (0.0000) | 0.0008 (0.0000) | 0.0008 (0.0000) | 0.0008 (0.0000) | 0.0008 (0.0000) | 0.0008 (0.0000) | 0.487 |
| Cingulum Cingulate Gyrus MD | 0.0008 (0.0000) | 0.0008 (0.0000) | 0.0008 (0.0000) | 0.0008 (0.0000) | 0.0008 (0.0000) | 0.0008 (0.0000) | 0.0008 (0.0000) | 0.018 |
| Cingulum Hippocampus MD | 0.0008 (0.0000) | 0.0008 (0.0000) | 0.0008 (0.0000) | 0.0008 (0.0000) | 0.0008 (0.0000) | 0.0008 (0.0000) | 0.0008 (0.0000) | <0.001 |
| Fornix Cresstria Terminalis MD | 0.0008 (0.0000) | 0.0008 (0.0000) | 0.0008 (0.0000) | 0.0008 (0.0000) | 0.0008 (0.0000) | 0.0008 (0.0000) | 0.0008 (0.0000) | 0.530 |
| Superior Longitudinal Fasciculus MD | 0.0007 (0.0000) | 0.0007 (0.0000) | 0.0007 (0.0000) | 0.0007 (0.0000) | 0.0007 (0.0000) | 0.0007 (0.0000) | 0.0007 (0.0000) | 0.087 |
| Superior Frontooccipital Fasciculus MD | 0.0007 (0.0001) | 0.0007 (0.0001) | 0.0007 (0.0001) | 0.0007 (0.0001) | 0.0007 (0.0001) | 0.0007 (0.0001) | 0.0007 (0.0001) | 0.526 |
| Uncinate Fasciculus MD | 0.0007 (0.0000) | 0.0007 (0.0000) | 0.0007 (0.0000) | 0.0007 (0.0000) | 0.0007 (0.0000) | 0.0007 (0.0000) | 0.0007 (0.0000) | 0.242 |
| Tapetum MD | 0.0010 (0.0001) | 0.0010 (0.0001) | 0.0010 (0.0001) | 0.0010 (0.0001) | 0.0010 (0.0001) | 0.0010 (0.0001) | 0.0010 (0.0001) | 0.268 |
| ^1^Mean (SD) | | | | | | | | |
| ^2^Kruskal-Wallis rank sum test | | | | | | | | |

*.

#### Supplementary Table 7. Association Between APOE Dosage (e4 and e2) and Neuroimaging Outcomes

See STable7_Fully_APOE4_and2_MRI.csv

#### Supplementary Table 8. Interaction Analysis of APOE Dosage and Sex on Neuroimaging Outcomes

See STable8_Fully_APOEXage_MRI.csv

#### Supplementary Table 9. Interaction Analysis of APOE Dosage and Sex on Neuroimaging Outcomes

See STable9_Fully_APOEXsex_MRI.csv

#### Supplementary Table 10. Association Between APOE Genotype (Categorical) and Neuroimaging Outcomes

See STable10_APOE_genotype_MRI.csv

#### Supplementary Table 11. Association Between Lifestyle Factors (Total and Component Scores) and Neuroimaging Outcomes

See STable11_Fully_Lifestyle_MRI.csv

#### Supplementary Table 12. Interaction Analysis of APOE Dosage and Total Lifestyle Score (Continuous) on Neuroimaging Outcomes

See STable12_Int_APOE_TLS_MRI.csv

#### Supplementary Table 13. Interaction Analysis of APOE Dosage and Total Lifestyle Score (Categorical) on Neuroimaging Outcomes

See STable13_APOE_catgorizeTLS.csv

#### Supplementary Table 14. Interaction Analysis of APOE Dosage and Lifestyle components (Binary) on Neuroimaging Outcomes

See STable14_APOE_subLS.csv

#### Supplementary Table 15. Interaction Analysis of APOE Genotype (Categorical) and Lifestyle Factors (Total and Component Scores) on Neuroimaging Outcomes

See STable15_APOE_genotype_allLS.csv

#### Supplementary Table 16. Interaction Analysis of APOE Genotype (Categorical) and Total Lifestyle Score (Categorical) on Neuroimaging Outcomes

See STable16_APOE_genotype_LS(CAT).csv

### Supplementary Figures

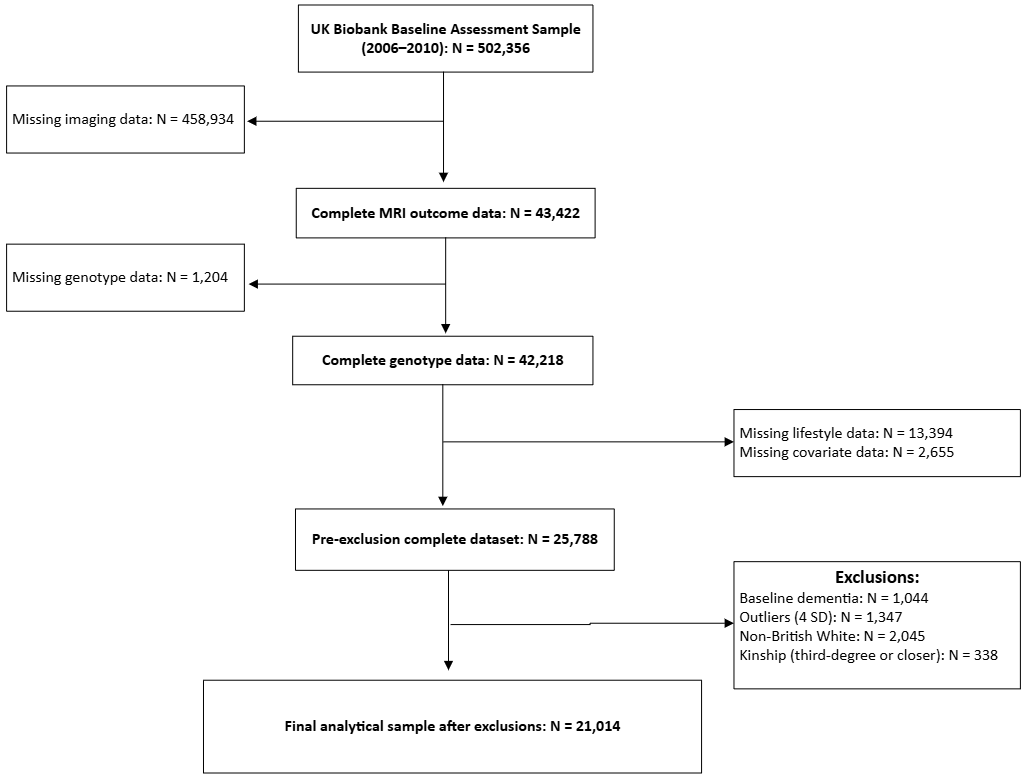

#### Supplementary Figure 1. The study population selection process

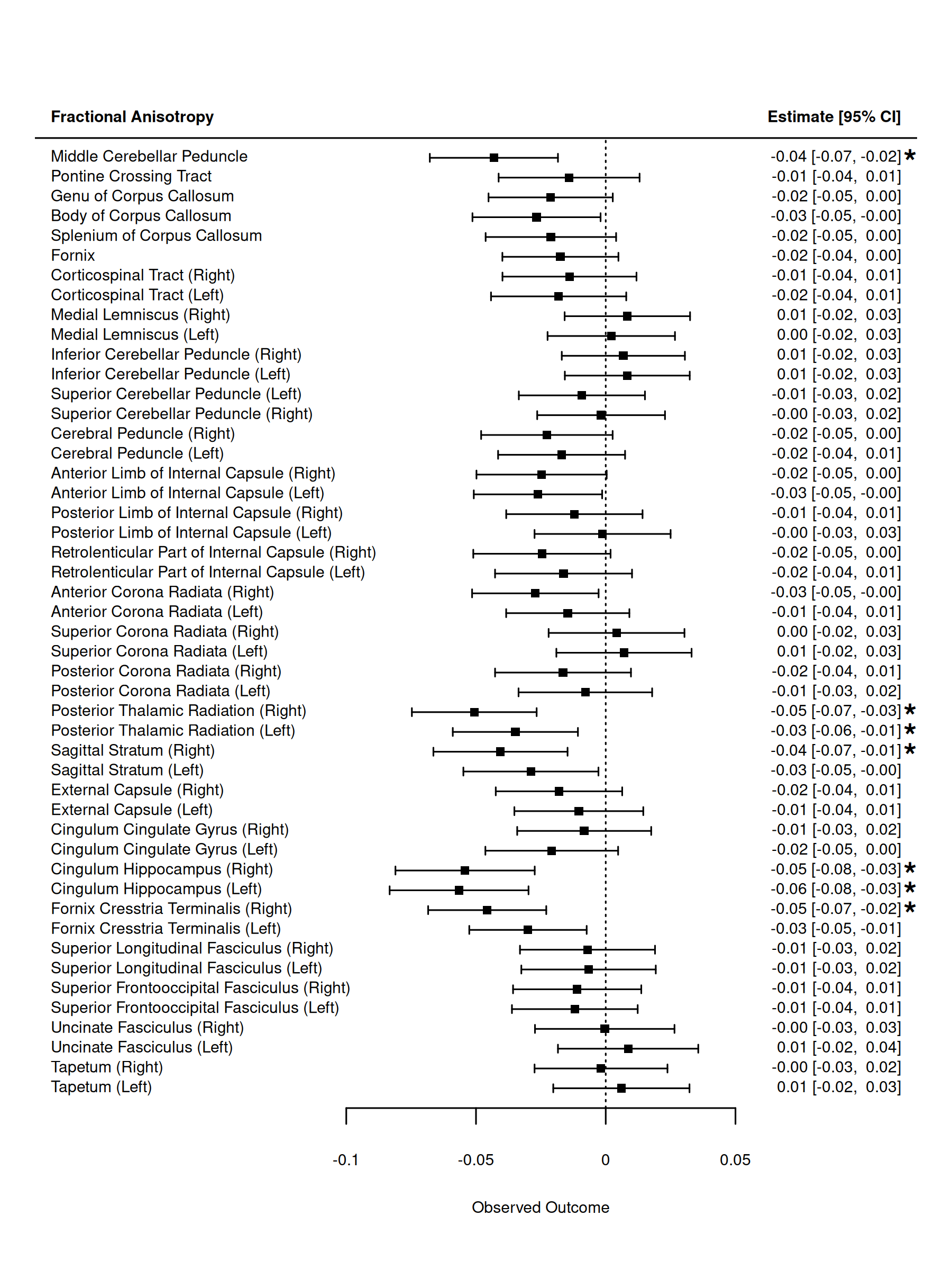

#### Supplementary Figure 2. Forest plots showing associations between APOE ε4 allele dosage and fractional anisotropy (FA) across white matter tracts.

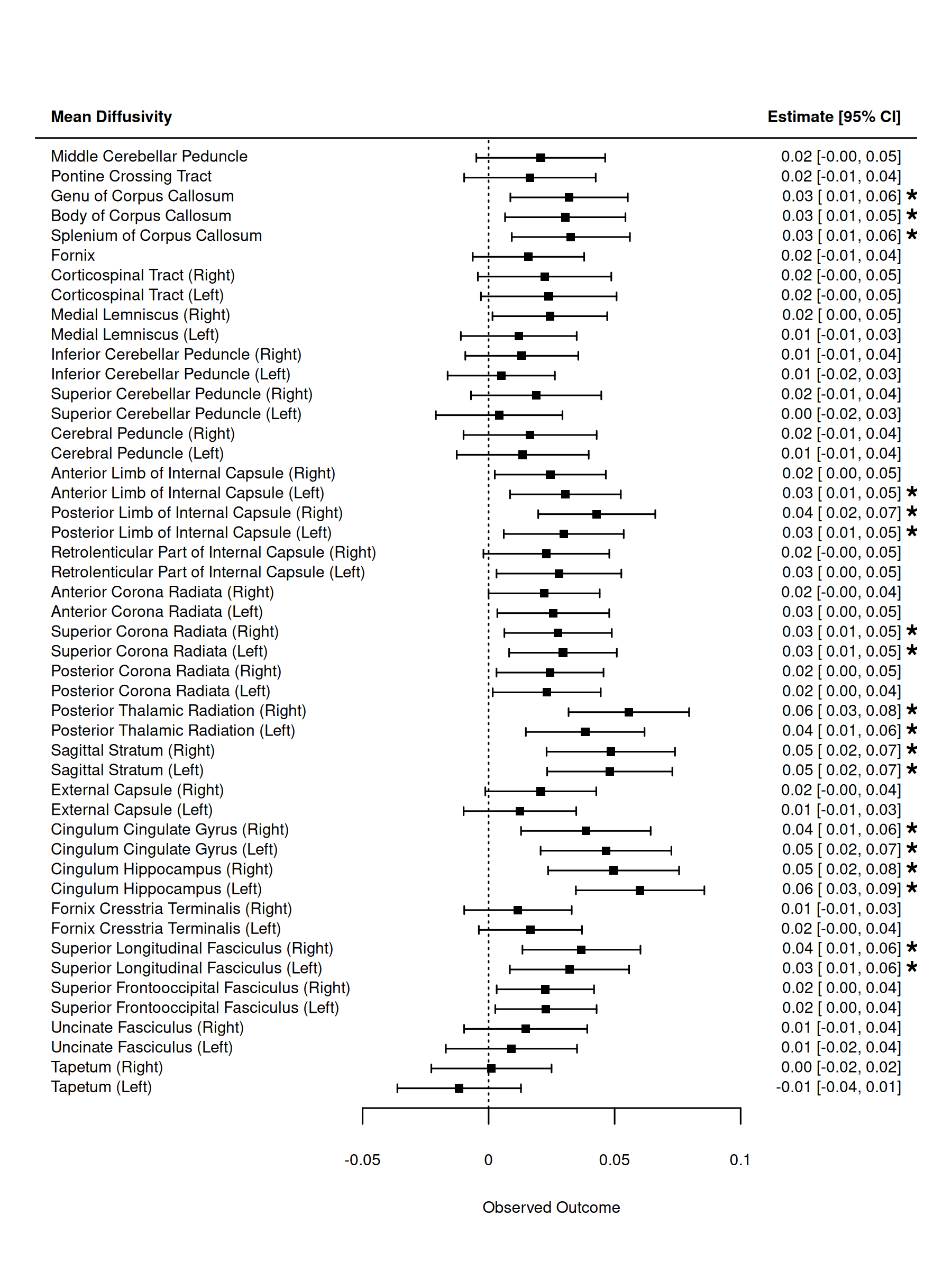

#### Supplementary Figure 3. Forest plots showing associations between APOE ε4 allele dosage and mean diffusivity (MD) across white matter tracts.
